## Supplemental file for "Species-specific detection of *Schistosoma japonicum* using the ‘SNAILS’ DNA-based biosensor"

^1^Section of Structural and Synthetic Biology, Department of Infectious Disease, Imperial College London, London, UK. ^2^Department of Parasitology, School of Basic Medical Sciences, Wuhan University, Wuhan 430071, Hubei Province, People’s Republic of China. ^3^Natural History Museum, London, UK. ^4^London School of Hygiene and Tropical Medicine, London, UK. ^5^The London Biofoundry, Imperial College Translation and Innovation Hub, White City Campus, London, UK. ^6^UK Dementia Research Institute Care Research and Technology Centre, Imperial College London, Hammersmith Campus, London, UK.

```
JQ781211.1      TGAATGATGGTTCCTTCAATATTTTATATGGAATTAAGTTTGTATTATGGATCTGGTGTT
KU196343.1      TGAATGATGGTTCCTTCAATATTTTATATGGAATTAAGTTTGTATTATGGATCTGGTGTT
KU196317.1      TGAATGATGGTTCCTTCAATATTTTATATGGAATTAAGTTTGTATTATGGATCTGGTGTT
HM120846.1      TGAATGATGGTTCCTTCAATATTTTATATGGAATTAAGTTTGTATTATGGATCTGGTGTT
KU196374.1      TGAATGATGGTTCCTTCAATATTTTATATGGAATTAAGTTTGTATTATGGATCTGGTGTT
KU196341.1      TGAATGATGGTTCCTTCAATATTTTATATGGAATTAAGTTTGTATTATGGATCTGGTGTT
KU196321.1      TGAATGATGGTTCCTTCAATATTTTATATGGAATTAAGTTTGTATTATGGATCTGGTGTT
KU196301.1      TGAATGATGGTTCCTTCAATATTTTATATGGAATTAAGTTTGTATTATGGATCTGGTGTT
KU196331.1      TGAATGATGGTTCCTTCAATATTTTATATGGAATTAAGTTTGTATTATGGATCTGGTGTT
KU196322.1      TGAATGATGGTTCCTTCAATATTTTATATGGAATTAAGTTTGTATTATGGATCTGGTGTT
KU196312.1      TGAATGATGGTTCCTTCAATATTTTATATGGAATTAAGTTTGTATTATGGATCTGGTGTT
EU325878.1      TGAATGATGGTTCCTTCAATATTTTATATGGAATTAAGTTTGTATTATGGATCTGGTGTT
KU196395.1      TGAATGATGGTTCCTTCAATATTTTATATGGAATTAAGTTTGTATTATGGATCTGGTGTT
KF279409.1      TGAATGATGGTTCCTTCAATATTTTATATGGAATTAAGTTTGTATTATGGATCTGGTGTT
KF279408.1      TGAATGATGGTTCCTTCAATATTTTATATGGAATTAAGTTTGTATTATGGATCTGGTGTT
KF279407.1      TGAATGATGGTTCCTTCAATATTTTATATGGAATTAAGTTTGTATTATGGATCTGGTGTT
HM120847.1      TGAATGATGGTTCCTTCAATATTTTATATGGAATTAAGTTTGTATTATGGATCTGGTGTT
KR855674.1      TGAATGATGGTTCCTTCAATATTTTATATGGAATTAAGTTTGTATTATGGATCTGGTGTT
KU196417.1      TGAATGATGGTTCCTTCAATATTTTATATGGAATTAAGTTTGTATTATGGATCTGGTGTT
KU196413.1      TGAATGATGGTTCCTTCAATATTTTATATGGAATTAAGTTTGTATTATGGATCTGGTGTT
KU196367.1      TGAATGATGGTTCCTTCAATATTTTATATGGAATTAAGTTTGTATTATGGATCTGGTGTT
HM120848.1      TGAATGATGGTTCCTTCAATATTTTATATGGAATTAAGTTTGTATTATGGATCTGGTGTT
JQ781208.1      TGAATGATGGTTCCTTCAATATTTTATATGGAATTAAGTTTGTATTATGGATCTGGTGTT
JQ781209.1      TGAATGATGGTTCCTTCAATATTTTATATGGAATTAAGTTTGTATTATGGATCTGGTGTT
JQ781213.1      TGAATGATGGTTCCTTCAATATTTTATATGGAATTAAGTTTGTATTATGGATCTGGTGTT
JQ781212.1      TGAATGATGGTTCCTTCAATATTTTATATGGAATTAAGTTTGTATTATGGATCTGGTGTT
JQ781206.1      TGAATGATGGTTCCTTCAATATTTTATATGGAATTAAGTTTGTATTATGGATCTGGTGTT
KU196355.1      TGAATGATGGTTCCTTCAATATTTTATATGGAATTAAGTTTGTATTATGGATCTGGTGTT
JQ781210.1      TGAATGATGGTTCCTTCAATATTTTATATGGAATTAAGTTTGTATTATGGATCTGGTGTT
KU196388.1      TGAATGATGGTTCCTTCAATATTTTATATGGAATTAAGTTTGTATTATGGATCTGGTGTT
KU196379.1      TGAATGATGGTTCCTTCAATATTTTATATGGAATTAAGTTTGTATTATGGATCTGGTGTT
JQ781214.1      TGAATGATGGTTCCTTCAATATTTTATATGGAATTAAGTTTGTATTATGGGTCTGGTGTT
KU196305.1      TGAATGATGGTTCCTTCAATATTTTATATGGAATTAAGTTTGTATTATGGATCTGGTGTT
                ************************************************** *********

JQ781211.1      GGTTGGACCTTTTATCCACCTTTGTCTTCTTTAGCTACTTCTGGTGTTGGTGTGGATTAC
KU196343.1      GGTTGGACTTTTTATCCACCTTTGTCTTCTTTAGCTACTTCTGGTGTTGGTGTGGATTAC
KU196317.1      GGTTGGACCTTTTATCCACCTTTGTCTTCTTTAGCTACTTCTGGTGTTGGTGTGGATTAC
HM120846.1      GGTTGGACCTTTTATCCACCTTTGTCTTCTTTAGCTACTTCTGGTGTTGGTGTGGATTAC
KU196374.1      GGTTGGACCTTTTATCCACCTTTGTCTTCTTTAGCTACTTCTGGTGTTGGTGTGGATTAC
KU196341.1      GGTTGGACCTTTTATCCACCTTTGTCTTCTTTAGCTACTTCTGGTGTTGGTGTGGATTAC
KU196321.1      GGTTGGACCTTTTATCCACCTTTGTCTTCTTTAGCTACTTCTGGTGTTGGTGTGGATTAC
KU196301.1      GGTTGGACCTTTTATCCACCTTTGTCTTCTTTAGCTACTTCTGGTGTTGGTGTGGATTAC
KU196331.1      GGTTGGACCTTTTATCCACCTTTGTCTTCTTTAGCTACTTCTGGTGTTGGTGTGGATTAC
KU196322.1      GGTTGGACCTTTTATCCACCTTTGTCTTCTTTAGCTACTTCTGGTGTTGGTGTGGATTAC
KU196312.1      GGTTGGACCTTTTATCCACCTTTGTCTTCTTTAGCTACTTCTGGTGTTGGTGTGGATTAC
EU325878.1      GGTTGGACCTTTTATCCACCTTTGTCTTCTTTAGCTACTTCTGGTGTTGGTGTGGATTAC
KU196395.1      GGTTGGACCTTTTATCCACCTTTGTCTTCTTTAGCTACTTCTGGTGTTGGTGTGGATTAC
KF279409.1      GGTTGGACCTTTTATCCACCTTTGTCTTCTTTAGCTACTTCTGGTGTTGGTGTGGATTAC
KF279408.1      GGTTGGACCTTTTATCCACCTTTGTCTTCTTTAGCTACTTCTGGTGTTGGTGTGGATTAC
KF279407.1      GGTTGGACCTTTTATCCACCTTTGTCTTCTTTAGCTACTTCTGGTGTTGGTGTGGATTAC
HM120847.1      GGTTGGACCTTTTATCCACCTTTGTCTTCTTTAGCTACTTCTGGTGTTGGTGTAGATTAC
KR855674.1      GGTTGGACCTTTTATCCACCTTTGTCTTCTTTAGCTACTTCTGGTGTTGGTGTGGATTAC
KU196417.1      GGTTGGACCTTTTATCCACCTTTGTCTTCTTTAGCTACTTCTGGTGTTGGTGTGGATTAC
KU196413.1      GGTTGGACCTTTTATCCACCTTTGTCTTCTTTAGCTACTTCTGGTGTTGGTGTGGATTAC
KU196367.1      GGTTGGACCTTTTATCCACCTTTGTCTTCTTTAGCTACTTCTGGTGTTGGTGTGGATTAC
HM120848.1      GGTTGGACCTTTTATCCACCTTTGTCTTCTTTAGCTACTTCTGGTGTTGGTGTGGATTAC
JQ781208.1      GGTTGGACCTTTTATCCACCTTTGTCTTCTTTAGCTACTTCTGGTGTTGGTGTGGATTAC
JQ781209.1      GGTTGGACCTTTTATCCACCTTTGTCTTCTTTAGCTACTTCTGGTGTTGGTGTGGATTAC
JQ781213.1      GGTTGGGCCTTTTATCCACCTTTGTCTTCTTTAGCTACTTCTGGTGTTGGTGTGGATTAC
JQ781212.1      GGTTGGACCTTTTATCCACCTTTGTCTTCTTTAGCTACTTCTGGTGTTGGTGTGGATTAC
JQ781206.1      GGTTGGACCTTTTATCCACCTTTGTCTTCTTTAGCTACTTCTGGTGTTGGTGTGGATTAC
KU196355.1      GGTTGGACCTTTTATCCACCTTTGTCTTCTTTAGCTACTTCTGGTGTTGGTGTGGATTAC
JQ781210.1      GGTTGGACCTTTTATCCACCTTTGTCTTCTTTAGCTACTTCTGGTGTTGGTGTGGATTAC
KU196388.1      GGTTGGACTTTTTATCCACCTTTGTCTTCTTTAGCTACTTCTGGTGTTGGTGTGGATTAC
KU196379.1      GGTTGGACTTTTTATCCACCTTTGTCTTCTTTAGCTACTTCTGGTGTTGGTGTGGATTAC
JQ781214.1      GGTTGGACCTTTTATCCACCTTTGTCTTCTTTAGCTACTTCTGGTGTTGGTGTGGATTAC
KU196305.1      GGTTGGACCTTTTATCCACCTTTGTCTTCTTTAGCTACTTCTGGTGTTGGTGTGGATTAC
                ****** * ******************************************** ******

JQ781211.1      TTAATGTTCTCTTTACATCTTGCTGGTGTATCTAGTTTGATTGGTTCTATAAATTTTATT
KU196343.1      TTAATGTTCTCTTTACATCTTGCTGGTGTATCTAGTTTGATTGGTTCTATAAATTTTATT
KU196317.1      TTAATGTTCTCTTTACATCTTGCTGGTGTATCTAGTTTGATTGGTTCTATAAATTTTATT
HM120846.1      TTAATGTTCTCTTTACATCTTGCTGGTGTATCTAGTTTGATTGGTTCTATAAATTTTATT
KU196374.1      TTAATGTTCTCTTTACATCTTGCTGGTGTATCTAGTTTGATTGGTTCTATAAATTTTATT
KU196341.1      TTAATGTTCTCTTTACATCTTGCTGGTGTATCTAGTTTGATTGGTTCTATAAATTTTATT
KU196321.1      TTAATGTTCTCTTTACATCTTGCTGGTGTATCTAGTTTGATTGGTTCTATAAATTTTATT
KU196301.1      TTAATGTTCTCTTTACATCTTGCTGGTGTATCTAGTTTGATTGGTTCTATAAATTTTATT
KU196331.1      TTAATGTTCTCTTTACATCTTGCTGGTGTATCTAGTTTGATTGGTTCTATAAATTTTATT
KU196322.1      TTAATGTTCTCTTTACATCTTGCTGGTGTATCTAGTTTGATTGGTTCTATAAATTTTATT
KU196312.1      TTAATGTTCTCTTTACATCTTGCTGGTGTATCTAGTTTGATTGGTTCTATAAATTTTATT
EU325878.1      TTAATGTTCTCTTTACATCTTGCTGGTGTATCTAGTTTGATTGGTTCTATAAATTTTATT
KU196395.1      TTAATGTTCTCTTTACATCTTGCTGGTGTATCTAGTTTGATTGGTTCTATAAATTTTATT
KF279409.1      TTAATGTTCTCTTTACATCTTGCTGGTGTATCTAGTTTGATTGGTTCTATAAATTTTATT
KF279408.1      TTAATGTTCTCTTTACATCTTGCTGGTGTATCTAGTTTGATTGGTTCTATAAATTTTATT
KF279407.1      TTAATGTTCTCTTTACATCTTGCTGGTGTATCTAGTTTGATTGGTTCTATAAATTTTATT
HM120847.1      TTAATGTTCTCTTTACATCTTGCTGGTGTATCTAGTTTGATTGGTTCTATAAATTTTATT
KR855674.1      TTAATGTTCTCTTTACATCTTGCTGGTGTATCTAGTTTGATTGGTTCTATAAATTTTATT
KU196417.1      TTAATGTTCTCTTTACATCTTGCTGGTGTATCTAGTTTGATTGGTTCTATAAATTTTATT
KU196413.1      TTAATGTTCTCTTTACATCTTGCTGGTGTATCTAGTTTGATTGGTTCTATAAATTTTATT
KU196367.1      TTAATGTTCTCTTTACATCTTGCTGGTGTATCTAGTTTGATTGGTTCTATAAATTTTATT
HM120848.1      TTAATGTTCTCTTTACATCTTGCTGGTGTATCTAGTTTGATTGGTTCTATAAATTTTATT
JQ781208.1      TTAATGTTCTCTTTACATCTTGCTGGTGTATCTAGTTTGATTGGTTCTATAAATTTTATT
JQ781209.1      TTAATGTTCTCTTTACATCTTGCTGGTGTATCTAGTTTGATTGGTTCTATAAATTTTATT
JQ781213.1      TTAATGTTCTCTTTACATCTTGCTGGTGTATCTAGTTTGATTGGTTCTATAAATTTTATT
JQ781212.1      TTAATGTTCTCTTTACATCTTGCTGGTGTATCTAGTTTGATTGGTTCTATAAATTTTATT
JQ781206.1      TTAATGTTCTCTTTACATCTTGCTGGTGTATCTAGTTTGATTGGTTCTATAAATTTTATT
KU196355.1      TTAATGTTCTCTTTACATCTTGCTGGTGTATCTAGTTTGATTGGTTCTATAAATTTTATT
JQ781210.1      TTAATGTTCTCTTTACATCTTGCTGGTGTATCTAGTTTGATTGGTTCTATAAATTTTATT
KU196388.1      TTAATGTTCTCTTTACATCTTGCTGGTGTATCTAGTTTGATTGGTTCTATAAATTTTATT
KU196379.1      TTAATGTTCTCTTTACATCTTGCTGGTGTATCTAGTTTGATTGGTTCTATAAATTTTATT
JQ781214.1      TTAATGTTCTCTTTACATCTTGCTGGTGTATCTAGTTTGATTGGTTCTATAAATTTTATT
KU196305.1      TTAATGTTCTCTTTACATCTTGCTGGTGTATCTAGTTTGATTGGTTCTATAAATTTTATT
                ************************************************************

JQ781211.1      ACTACTATAATGTTGCGTCTAAGGTCATGCTCTTCAGTTATTAGATGATCTTATTTATTT
KU196343.1      ACTACTATAATGTTGCGTCTAAGGTCATGTTCTTCAGTTATTAGATGATCTTATTTATTT
KU196317.1      ACTACTATAATGTTGCGTCTAAGGTCATGTTCTTCAGTTATTAGATGATCTTATTTATTT
HM120846.1      ACTACTATAATGTTGCGTCTAAGGTCATGTTCTTCAGTTATTAGATGATCTTATTTATTT
KU196374.1      ACTACTATAATGTTGCGTCTAAGGTCATGTTCTTCAGTTATTAGATGATCTTATTTATTT
KU196341.1      ACTACTATAATGTTGCGTCTAAGGTCATGTTCTTCAGTTATTAGATGATCTTATTTATTT
KU196321.1      ACTACTATAATGTTGCGTCTAAGGTCATGTTCTTCAGTTATTAGATGATCTTATTTATTT
KU196301.1      ACTACTATAATGTTGCGTCTAAGGTCATGTTCTTCAGTTATTAGATGATCTTATTTATTT
KU196331.1      ACTACTATAATGTTGCGTCTAAGGTCATGTTCTTCAGTTATTAGATGATCTTATTTATTT
KU196322.1      ACTACTATAATGTTGCGTCTAAGGTCATGTTCTTCAGTTATTAGATGATCTTATTTATTT
KU196312.1      ACTACTATAATGTTGCGTCTAAGGTCATGTTCTTCAGTTATTAGATGATCTTATTTATTT
EU325878.1      ACTACTATAATGTTGCGTCTAAGGTCATGTTCTTCAGTTATTAGATGATCTTATTTATTT
KU196395.1      ACTACTATAATGTTGCGTCTAAGGTCATGTTCTTCAGTTATTAGATGATCTTATTTATTT
KF279409.1      ACTACTATAATGTTGCGTCTAAGGTCATGTTCTTCAGTTATTAGATGATCTTATTTATTT
KF279408.1      ACTACTATAATGTTGCGTCTAAGGTCATGTTCTTCAGTTATTAGATGATCTTATTTATTT
KF279407.1      ACTACTATAATGTTGCGTCTAAGGTCATGTTCTTCAGTTATTAGATGATCTTATTTATTT
HM120847.1      ACTACTATAATGTTGCGTCTAAGGTCATGTTCTTCAGTTATTAGATGATCTTATTTATTT
KR855674.1      ACTACTATAATGTTGCGTCTAAGGTCATGTTCTTCAGTTATTAGATGATCTTATTTATTT
KU196417.1      ACTACTATAATGTTGCGTCTAAGGTCATGTTCTTCAGTTATTAGATGATCTTATTTATTT
KU196413.1      ACTACTATAATGTTGCGTCTAAGGTCATGTTCTTCAGTTATTAGATGATCTTATTTATTT
KU196367.1      ACTACTATAATGTTGCGTCTAAGGTCATGTTCTTCAGTTATTAGATGATCTTATTTATTT
HM120848.1      ACTACTATAATGTTGCGTCTAAGGTCATGTTCTTCAGTTATTAGATGATCTTATTTATTT
JQ781208.1      ACTACTATAATGTTGCGTCTAAGGTCATGTTCTTCAGTTATTAGATGATCTTATTTATTT
JQ781209.1      ACTACTATAATGTTGCGTCTAAGGTCATGTTCTTCAGTTATTAGATGATCTTATTTATTT
JQ781213.1      ACTACTATAATGTTGCGTCTAAGGTCATGTTCTTCAGTTATTAGATGATCTTATTTATTT
JQ781212.1      ACTACTATAATGTTGCGTCTAAGGTCATGTTCTTCAGTTATTAGATGATCTTATTTATTT
JQ781206.1      ACTACTATAATGTTGCGTCTAAGGTCATGTTCTTCAGTTATTAGATGATCTTATTTATTT
KU196355.1      ACTACTATAATGTTGCGTCTAAGGTCATGTTCTTCAGTTATTAGATGATCTTATTTATTT
JQ781210.1      ACTACTATAATGTTGCGTCTAAGGTCATGTTCTTCAGTTATTAGATGATCTTATTTATTT
KU196388.1      ACTACTATAATGTTGCGTCTAAGGTCATGTTCTTCAGTTATTAGATGATCTTATTTATTT
KU196379.1      ACTACTATAATGTTGCGTCTAAGGTCATGTTCTTCAGTTATTAGATGATCTTATTTATTT
JQ781214.1      ACTACTATAATGTTGCGTCTAAGGTCATGTTCTTCAGTTATTAGATGATCTTATTTATTT
KU196305.1      ACTACTATAATGTTGCGTCTAAGGTCATGTTCTTCAGTTATTAGATGATCTTATTTATTT
                ***************************** ******************************

JQ781211.1      ACTTCGGTGTTGTTATTGTTATCGTTGCCGGTTCTTGCTGCAGGTATAACTATGTTGTTG
KU196343.1      ACTTCGGTGTTGTTATTGTTATCGTTGCCGGTTCTTGCTGCAGGTATAACTATGTTGTTG
KU196317.1      ACTTCGGTGTTGTTATTGTTATCGTTGCCGGTTCTTGCTGCAGGTATAACTATGTTGTTG
HM120846.1      ACTTCGGTGTTGTTATTGTTATCGTTGCCGGTTCTTGCTGCAGGTATAACTATGTTGTTG
KU196374.1      ACTTCGGTGTTGTTATTGTTATCGTTGCCGGTTCTTGCTGCAGGTATAACTATGTTGTTG
KU196341.1      ACTTCGGTGTTGTTATTGTTATCGTTGCCGGTTCTTGCTGCAGGTATAACTATGTTGTTG
KU196321.1      ACTTCGGTGTTGTTATTGTTATCGTTGCCGGTTCTTGCTGCAGGTATAACTATGTTGTTG
KU196301.1      ACTTCGGTGTTGTTATTGTTATCGTTGCCGGTTCTTGCTGCAGGTATAACTATGTTGTTG
KU196331.1      ACTTCGGTGTTGTTATTGTTATCGTTGCCGGTTCTTGCTGCAGGTATAACTATGTTGTTG
KU196322.1      ACTTCGGTGTTGTTATTGTTATCGTTGCCGGTTCTTGCTGCAGGTATAACTATGTTGTTG
KU196312.1      ACTTCGGTGTTGTTATTGTTATCGTTGCCGGTTCTTGCTGCAGGTATAACTATGTTGTTG
EU325878.1      ACTTCGGTGTTGTTATTGTTATCGTTGCCGGTTCTTGCTGCAGGTATAACTATGTTGTTG
KU196395.1      ACTTCGGTGTTGTTATTGTTATCGTTGCCGGTTCTTGCTGCAGGTATAACTATGTTGTTG
KF279409.1      ACTTCGGTGTTGTTATTGTTATCGTTGCCGGTTCTTGCTGCAGGTATAACTATGTTGTTG
KF279408.1      ACTTCGGTGTTGTTATTGTTATCGTTGCCGGTTCTTGCTGCAGGTATAACTATGTTGTTG
KF279407.1      ACTTCGGTGTTGTTATTGTTATCGTTGCCGGTTCTTGCTGCAGGTATAACTATGTTGTTG
HM120847.1      ACTTCGGTGTTGTTATTGTTATCGTTGCCGGTTCTTGCTGCAGGTATAACTATGTTGTTG
KR855674.1      ACTTCGGTGTTGTTATTGTTATCGTTGCCGGTTCTTGCTGCAGGTATAACTATGTTGTTG
KU196417.1      ACTTCGGTGTTGTTATTGTTATCGTTGCCGGTTCTTGCTGCAGGTATAACTATGTTGTTG
KU196413.1      ACTTCGGTGTTGTTATTGTTATCGTTGCCGGTTCTTGCTGCAGGTATAACTATGTTGTTG
KU196367.1      ACTTCGGTGTTGTTATTGTTATCGTTGCCGGTTCTTGCTGCAGGTATAACTATGTTGTTG
HM120848.1      ACTTCGGTGTTGTTATTGTTATCGTTGCCGGTTCTTGCTGCAGGTATAACTATGTTGTTG
JQ781208.1      ACTTCGGTGTTGTTATTGTTATCGTTGCCGGTTCTTGCTGCAGGTATAACTATGTTGTTG
JQ781209.1      ACTTCGGTGTTGTTATTGTTATCGTTGCCGGTTCTTGCTGCAGGTATAACTATGTTGTTG
JQ781213.1      ACTTCGGTGTTGTTATTGTTATCGTTGCCGGTTCTTGCTGCAGGTATAACTATGTTGTTG
JQ781212.1      ACTTCGGTGTTGTTATTGTTATCGTTGCCGGTTCTTGCTGCAGGTATAACTATGTTGTTG
JQ781206.1      ACTTCGGTGTTGTTATTGTTATCGTTGCCGGTTCTTGCTGCAGGTATAACTATGTTGTTG
KU196355.1      ACTTCGGTGTTGTTATTGTTATCGTTGCCGGTTCTTGCTGCAGGTATAACTATGTTGTTG
JQ781210.1      ACTTCGGTGTTGTTATTGTTATCGTTGCCGGTTCTTGCTGCAGGTATAACTATGTTGTTG
KU196388.1      ACTTCGGTGTTGTTATTGTTATCGTTGCCGGTTCTTGCTGCAGGTATAACTATGTTGTTG
KU196379.1      ACTTCGGTGTTGTTATTGTTATCGTTGCCGGTTCTTGCTGCAGGTATAACTATGTTGTTG
JQ781214.1      ACTTCGGTGTTGTTATTGTTATCGTTGCCGGTTCTTGCTGCAGGTATAACTATGTTGTTG
KU196305.1      ACTTCGGTGTTGTTATTGTTATCGTTGCCGGTTCTTGCTGCAGGTATAACTATGTTGTTG
                ************************************************************

JQ781211.1      TTTGATCGTAAATTTGGTACTGCTTTTTTTGAGCCAGCAGGTGGTGGTGATCCTGTGTTA
KU196343.1      TTTGATCGTAAATTTGGTACTGCTTTTTTTGAGCCAGCAGGTGGTGGTGATCCTGTGTTA
KU196317.1      TTTGATCGTAAATTTGGTACTGCTTTTTTTGAGCCAGCAGGTGGTGGTGATCCTGTGTTA
HM120846.1      TTTGATCGTAAATTTGGTACTGCTTTTTTTGAGCCAGCAGGTGGTGGTGATCCTGTGTTA
KU196374.1      TTTGATCGTAAATTTGGTACTGCTTTTTTTGAGCCAGCAGGTGGTGGTGATCCTGTGTTA
KU196341.1      TTTGATCGTAAATTTGGTACTGCTTTTTTTGAGCCAGCAGGTGGTGGTGATCCTGTGTTA
KU196321.1      TTTGATCGTAAATTTGGTACTGCTTTTTTTGAGCCAGCAGGTGGTGGTGATCCTGTGTTA
KU196301.1      TTTGATCGTAAATTTGGTACTGCTTTTTTTGAGCCAGCAGGTGGTGGTGATCCTGTGTTA
KU196331.1      TTTGATCGTAAATTTGGTACTGCTTTTTTTGAGCCAGCAGGTGGTGGTGATCCTGTGTTA
KU196322.1      TTTGATCGTAAATTTGGTACTGCTTTTTTTGAGCCAGCAGGTGGTGGTGATCCTGTGTTA
KU196312.1      TTTGATCGTAAATTTGGTACTGCTTTTTTTGAGCCAGCAGGTGGTGGTGATCCTGTGTTA
EU325878.1      TTTGATCGTAAATTTGGTACTGCTTTTTTTGAGCCAGCAGGTGGTGGTGATCCTGTGTTA
KU196395.1      TTTGATCGTAAATTTGGTACTGCTTTTTTTGAGCCAGCAGGTGGTGGTGATCCTGTGTTA
KF279409.1      TTTGATCGTAAATTTGGTACTGCTTTTTTTGAGCCAGCAGGTGGTGGTGATCCTGTGTTA
KF279408.1      TTTGATCGTAAATTTGGTACTGCTTTTTTTGAGCCAGCAGGTGGTGGTGATCCTGTGTTA
KF279407.1      TTTGATCGTAAATTTGGTACTGCTTTTTTTGAGCCAGCAGGTGGTGGTGATCCTGTGTTA
HM120847.1      TTTGATCGTAAATTTGGTACTGCTTTTTTTGAGCCAGCAGGTGGTGGTGATCCTGTGTTA
KR855674.1      TTTGATCGTAAATTTGGTACTGCTTTTTTTGAGCCAGCAGGTGGTGGTGATCCTGTGTTA
KU196417.1      TTTGATCGTAAATTTGGTACTGCTTTTTTTGAGCCAGCAGGTGGTGGTGATCCTGTGTTA
KU196413.1      TTTGATCGTAAATTTGGTACTGCTTTTTTTGAGCCAGCAGGTGGTGGTGATCCTGTGTTA
KU196367.1      TTTGATCGTAAATTTGGTACTGCTTTTTTTGAGCCAGCAGGTGGTGGTGATCCTGTGTTA
HM120848.1      TTTGATCGTAAATTTGGTACTGCTTTTTTTGAGCCAGCAGGTGGTGGTGATCCTGTGTTA
JQ781208.1      TTTGATCGTAAATTTGGTACTGCTTTTTTTGAGCCAGCAGGTGGTGGTGATCCTGTGTTA
JQ781209.1      TTTGATCGTAAATTTGGTACTGCTTTTTTTGAGCCAGCAGGTGGTGGTGATCCTGTGTTA
JQ781213.1      TTTGATCGTAAATTTGGTACTGCTTTTTTTGAGCCAGCAGGTGGTGGTGATCCTGTGTTA
JQ781212.1      TTTGATCGTAAATTTGGTACTGCTTTTTTTGAGCCAGCAGGTGGTGGTGATCCTGTGTTA
JQ781206.1      TTTGATCGTAAATTTGGTACTGCTTTTTTTGAGCCAGCAGGTGGTGGTGATCCTGTGTTA
KU196355.1      TTTGATCGTAAATTTGGTACTGCTTTTTTTGAGCCAGCAGGTGGTGGTGATCCTGTGTTA
JQ781210.1      TTTGATCGTAAATTTGGTACTGCTTTTTTTGAGCCAGCAGGTGGTGGTGATCCTGTGTTA
KU196388.1      TTTGATCGTAAATTTGGTACTGCTTTTTTTGAGCCAGCAGGTGGTGGTGATCCTGTGTTA
KU196379.1      TTTGATCGTAAATTTGGTACTGCTTTTTTTGAGCCAGCAGGTGGTGGTGATCCTGTGTTA
JQ781214.1      TTTGATCGTAAATTTGGTACTGCTTTTTTTGAGCCAGCAGGTGGTGGTGATCCTGTGTTA
KU196305.1      TTTGATCGTAAATTTGGTACTGCTTTTTTTGAGCCAGCAGGTGGTGGTGATCCTGTGTTA
                ************************************************************

JQ781211.1      TTTCAACATTTATTTTGGTTTTTTGGTCACCCAGAAGTATATGTTTTGATATTGCCTGGA
KU196343.1      TTTCAACATTTATTTTGGTTTTTTGGTCACCCAGAAGTATATGTTTTGATATTGCCTGGA
KU196317.1      TTTCAACATTTATTTTGGTTTTTTGGTCACCCAGAAGTATATGTTTTGATATTGCCTGGA
HM120846.1      TTTCAACATTTATTTTGGTTTTTTGGTCACCCAGAAGTATATGTTTTGATATTGCCTGGA
KU196374.1      TTTCAACATTTATTTTGGTTTTTTGGTCACCCAGAAGTATATGTTTTGATATTGCCTGGA
KU196341.1      TTTCAACATTTATTTTGGTTTTTTGGTCACCCAGAAGTATATGTTTTGATATTGCCTGGA
KU196321.1      TTTCAACATTTATTTTGGTTTTTTGGTCACCCAGAAGTATATGTTTTGATATTGCCTGGA
KU196301.1      TTTCAACATTTATTTTGGTTTTTTGGTCACCCAGAAGTATATGTTTTGATATTGCCTGGA
KU196331.1      TTTCAACATTTATTTTGGTTTTTTGGTCACCCAGAAGTATATGTTTTGATATTGCCTGGA
KU196322.1      TTTCAACATTTATTTTGGTTTTTTGGTCACCCAGAAGTATATGTTTTGATATTGCCTGGA
KU196312.1      TTTCAACATTTATTTTGGTTTTTTGGTCACCCAGAAGTATATGTTTTGATATTGCCTGGA
EU325878.1      TTTCAACATTTATTTTGGTTTTTTGGTCACCCAGAAGTATATGTTTTGATATTGCCTGGA
KU196395.1      TTTCAACATTTATTTTGGTTTTTTGGTCACCCAGAAGTATATGTTTTGATATTGCCTGGA
KF279409.1      TTTCAACATTTATTTTGGTTTTTTGGTCACCCAGAAGTATATGTTTTGATATTGCCTGGA
KF279408.1      TTTCAACATTTATTTTGGTTTTTTGGTCACCCAGAAGTATATGTTTTGATATTGCCTGGA
KF279407.1      TTTCAACATTTATTTTGGTTTTTTGGTCACCCAGAAGTATATGTTTTGATATTGCCTGGA
HM120847.1      TTTCAACATTTATTTTGGTTTTTTGGTCACCCAGAAGTATATGTTTTGATATTGCCTGGA
KR855674.1      TTTCAACATTTATTTTGGTTTTTTGGTCACCCAGAAGTATATGTTTTGATATTGCCTGGA
KU196417.1      TTTCAACATTTATTTTGGTTTTTTGGTCACCCAGAAGTATATGTTTTGATATTGCCTGGA
KU196413.1      TTTCAACATTTATTTTGGTTTTTTGGTCACCCAGAAGTATATGTTTTGATATTGCCTGGA
KU196367.1      TTTCAACATTTATTTTGGTTTTTTGGTCACCCAGAAGTATATGTTTTGATATTGCCTGGA
HM120848.1      TTTCAACATTTATTTTGGTTTTTTGGTCACCCAGAAGTATATGTTTTGATATTGCCTGGA
JQ781208.1      TTTCAACATTTATTTTGGTTTTTTGGTCACCCAGAAGTATATGTTTTGATATTGCCTGGA
JQ781209.1      TTTCAACATTTATTTTGGTTTTTTGGTCACCCAGAAGTATATGTTTTGATATTGCCTGGA
JQ781213.1      TTTCAACATTTATTTTGGTTTTTTGGTCACCCAGAAGTATATGTTTTGATATTGCCTGGA
JQ781212.1      TTTCAACATTTATTTTGGTTTTTTGGTCACCCAGAAGTATATGTTTTGATATTGCCTGGA
JQ781206.1      TTTCAACATTTATTTTGGTTTTTTGGTCACCCAGAAGTATATGTTTTGATATTGCCTGGA
KU196355.1      TTTCAACATTTATTTTGGTTTTTTGGTCACCCAGAAGTATATGTTTTGATATTGCCTGGA
JQ781210.1      TTTCAACATTTATTTTGGTTTTTTGGTCACCCAGAAGTATATGTTTTGATATTGCCTGGA
KU196388.1      TTTCAACATTTATTTTGGTTTTTTGGTCACCCAGAAGTATATGTTTTGATATTGCCTGGA
KU196379.1      TTTCAACATTTATTTTGGTTTTTTGGTCACCCAGAAGTATATGTTTTGATATTGCCTGGA
JQ781214.1      TTTCAACATTTATTTTGGTTTTTTGGTCACCCAGAAGTATATGTTTTGATATTGCCTGGA
KU196305.1      TTTCAACATTTATTTTGGTTTTTTGGTCACCCAGAAGTATATGTTTTGATATTGCCTGGA
                ************************************************************

JQ781211.1      TTTGGTATAGTAAGTCATATATGTATGTCTTTAAGTAATAATAATTCTTCGTTTGGATAT
KU196343.1      TTTGGTATAGTAAGTCATATATGTATGTCTTTAAGTAATAATAATTCTTCGTTTGGATAT
KU196317.1      TTTGGTATAGTAAGTCATATATGTATGTCTTTAAGTAATAATAATTCTTCGTTTGGATAT
HM120846.1      TTTGGTATAGTAAGTCATATATGTATGTCTTTAAGTAATAATAATTCTTCGTTTGGATAT
KU196374.1      TTTGGTATAGTAAGTCATATATGTATGTCTTTAAGTAATAATAATTCTTCGTTTGGATAT
KU196341.1      TTTGGTATAGTAAGTCATATATGTATGTCTTTAAGTAATAATAATTCTTCGTTTGGATAT
KU196321.1      TTTGGTATAGTAAGTCATATATGTATGTCTTTAAGTAATAATAATTCTTCGTTTGGATAT
KU196301.1      TTTGGTATAGTAAGTCATATATGTATGTCTTTAAGTAATAATAATTCTTCGTTTGGATAT
KU196331.1      TTTGGTATAGTAAGTCATATATGTATGTCTTTAAGTAATAATAATTCTTCGTTTGGATAT
KU196322.1      TTTGGTATAGTAAGTCATATATGTATGTCTTTAAGTAATAATAATTCTTCGTTTGGATAT
KU196312.1      TTTGGTATAGTAAGTCATATATGTATGTCTTTAAGTAATAATAATTCTTCGTTTGGATAT
EU325878.1      TTTGGTATAGTAAGTCATATATGTATGTCTTTAAGTAATAATAATTCTTCGTTTGGATAT
KU196395.1      TTTGGTATAGTAAGTCATATATGTATGTCTTTAAGTAATAATAATTCTTCGTTTGGATAT
KF279409.1      TTTGGTATAGTAAGTCATATATGTATGTCTTTAAGTAATAATAATTCTTCGTTTGGATAT
KF279408.1      TTTGGTATAGTAAGTCATATATGTATGTCTTTAAGTAATAATAATTCTTCGTTTGGATAT
KF279407.1      TTTGGTATAGTAAGTCATATATGTATGTCTTTAAGTAATAATAATTCTTCGTTTGGATAT
HM120847.1      TTTGGTATAGTAAGTCATATATGTATGTCTTTAAGTAATAATAATTCTTCGTTTGGATAT
KR855674.1      TTTGGTATAGTAAGTCATATATGTATGTCTTTAAGTAATAATAATTCTTCGTTTGGATAT
KU196417.1      TTTGGTATAGTAAGTCATATATGTATGTCTTTAAGTAATAATAATTCTTCGTTTGGATAT
KU196413.1      TTTGGTATAGTAAGTCATATATGTATGTCTTTAAGTAATAATAATTCTTCGTTTGGATAT
KU196367.1      TTTGGTATAGTAAGTCATATATGTATGTCTTTAAGTAATAATAATTCTTCGTTTGGATAT
HM120848.1      TTTGGTATAGTAAGTCATATATGTATGTCTTTAAGTAATAATAATTCTTCGTTTGGATAT
JQ781208.1      TTTGGTATAGTAAGTCATATATGTATGTCTTTAAGTAATAATAATTCTTCGTTTGGATAT
JQ781209.1      TTTGGTATAGTAAGTCATATATGTATGTCTTTAAGTAATAATAATTCTTCGTTTGGATAT
JQ781213.1      TTTGGTATAGTAAGTCATATATGTATGTCTTTAAGTAATAATAATTCTTCGTTTGGATAT
JQ781212.1      TTTGGTATAGTAAGTCATATATGTATGTCTTTAAGTAATAATAATTCTTCGTTTGGATAT
JQ781206.1      TTTGGTATAGTAAGTCATATATGTATGTCTTTAAGTAATAATAATTCTTCGTTTGGATAT
KU196355.1      TTTGGTATAGTAAGTCATATATGTATGTCTTTAAGTAATAATAATTCTTCGTTTGGATAT
JQ781210.1      TTTGGTATAGTAAGTCATATATGTATGTCTTTAAGTAATAATAATTCTTCGTTTGGATAT
KU196388.1      TTTGGTATAGTAAGTCATATATGTATGTCTTTAAGTAATAATAATTCTTCGTTTGGATAT
KU196379.1      TTTGGTATAGTAAGTCATATATGTATGTCTTTAAGTAATAATAATTCTTCGTTTGGATAT
JQ781214.1      TTTGGTATAGTAAGTCATATATGTATGTCTTTAAGTAATAATAATTCTTCGTTTGGATAT
KU196305.1      TTTGGTATAGTAAGTCATATATGTATGTCTTTAAGTAATAATAATTCTTCGTTTGGATAT
                ************************************************************

JQ781211.1      TGTGGGTTAGTTTGTGCTATGGGTTCTATTGTGTGTTTGGGGAGAGTTGTTTGGGCTCAT
KU196343.1      TATGGGTTAGTTTGTGCCATGGGTTCTATTGTGTGTTTGGGGAGAGTTGTTTGGGCTCAC
KU196317.1      TATGGGTTAGTTTGTGCTATGGGTTCTATTGTGTGTTTGGGGAGAGTTGTTTGGGCTCAC
HM120846.1      TATGGGTTAGTTTGTGCTATGGGTTCTATTGTGTGTTTGGGGAGAGTTGTTTGGGCTCAC
KU196374.1      TATGGGTTAGTTTGTGCTATGGGTTCTATTGTGTGTTTGGGGAGAGTTGTTTGGGCTCAC
KU196341.1      TATGGGTTAGTTTGTGCTATGGGTTCTATTGTGTGTTTGGGGAGAGTTGTTTGGGCTCAC
KU196321.1      TATGGGTTAGTTTGTGCTATGGGTTCTATTGTGTGTTTGGGGAGAGTTGTTTGGGCTCAC
KU196301.1      TATGGGTTAGTTTGTGCTATGGGTTCTATTGTGTGTTTGGGGAGAGTTGTTTGGGCTCAC
KU196331.1      TATGGGTTAGTTTGTGCTATGGGTTCTATTGTGTGTTTGGGGAGAGTTGTTTGGGCTCAC
KU196322.1      TATGGGTTAGTTTGTGCTATGGGTTCTATTGTGTGTTTGGGGAGAGTTGTTTGGGCTCAC
KU196312.1      TATGGGTTAGTTTGTGCTATGGGTTCTATTGTGTGTTTGGGGAGAGTTGTTTGGGCTCAC
EU325878.1      TATGGGTTAGTTTGTGCTATGGGTTCTATTGTGTGTTTGGGGAGAGTTGTTTGGGCTCAC
KU196395.1      TATGGGTTAGTTTGTGCTATGGGTTCTATTGTGTGTTTGGGGAGAGTTGTTTGGGCTCAC
KF279409.1      TATGGGTTAGTTTGTGCTATGGGTTCTATTGTGTGTTTGGGGAGAGTTGTTTGGGCTCAC
KF279408.1      TATGGGTTAGTTTGTGCTATGGGTTCTATTGTGTGTTTGGGGAGAGTTGTTTGGGCTCAC
KF279407.1      TATGGGTTAGTTTGTGCTATGGGTTCTATTGTGTGTTTGGGGAGAGTTGTTTGGGCTCAC
HM120847.1      TATGGGTTAGTTTGTGCTATGGGTTCTATTGTGTGTTTGGGGAGAGTTGTTTGGGCTCAC
KR855674.1      TATGGGTTAGTTTGTGCTATGGGTTCTATTGTGTGTTTGGGGAGAGTTGTTTGGGCTCAC
KU196417.1      TATGGGTTAGTTTGTGCTATGGGTTCTATTGTGTGTTTGGGGAGAGTTGTTTGGGCTCAC
KU196413.1      TATGGGTTAGTTTGTGCTATGGGTTCTATTGTGTGTTTGGGGAGAGTTGTTTGGGCTCAC
KU196367.1      TATGGGTTAGTTTGTGCTATGGGTTCTATTGTGTGTTTGGGGAGAGTTGTTTGGGCTCAC
HM120848.1      TATGGGTTAGTTTGTGCTATGGGTTCTATTGTGTGTTTGGGGAGAGTTGTTTGGGCTCAC
JQ781208.1      TATGGGTTAGTTTGTGCTATGGGTTCTATTGTGTGTTTGGGGAGAGTTGTTTGGGCTCAT
JQ781209.1      TATGGGTTAGTTTGTGCTATGGGTTCTATTGTGTGTTTGGGGAGAGTTGTTTGGGCTCAT
JQ781213.1      TATGGGTTAGTTTGTGCTATGGGTTCTATTGTGTGTTTGGGGAGAGTTGTTTGGGCTCAT
JQ781212.1      TATGGGTTAGTTTGTGCTATGGGTTCTATTGTGTGTTTGGGGAGAGTTGTTTGGGCTCAT
JQ781206.1      TATGGGTTAGTTTGTGCTATGGGTTCTATTGTGTGTTTGGGGAGAGTTGTTTGGGCTCAT
KU196355.1      TATGGGTTAGTTTGTGCTATGGGTTCTATTGTGTGTTTGGGGAGAGTTGTTTGGGCTCAT
JQ781210.1      TATGGGTTAGTTTGTGCTATGGGTTCTATTGTGTGTTTGGGGAGAGTTGTTTGGGCTCAT
KU196388.1      TATGGGTTAGTTTGTGCTATGGGTTCTATTGTGTGTTTGGGGAGAGTTGTTTGGGCTCAT
KU196379.1      TATGGGTTAGTTTGTGCTATGGGTTCTATTGTGTGTTTGGGGAGAGTTGTTTGGGCTCAT
JQ781214.1      TATGGGTTAGTTTGTGCTATGGGTTCTATTGTGTGTTTGGGGAGAGTTGTTTGGGCTCAT
KU196305.1      TATGGGTTAGTTTGTGCTATGGGTTCTATTGTGTGTTTGGGGAGAGTTGTTTGGGCTCAT
                * *************** *****************************************

JQ781211.1      CATATGTTTATGGTTGGTATGGATGTAAAGACTTCTGTTTTTTTTAGTTCTGTAACAATG
KU196343.1      CATATGTTTATGGTTGGTATGGATGTAAAGACTTCTGTTTTTTTTAGTTCTGTAACAATG
KU196317.1      CATATGTTTATGGTTGGTATGGATGTAAAGACTTCTGTTTTTTTTAGTTCTGTAACAATG
HM120846.1      CATATGTTTATGGTTGGTATGGATGTAAAGACTTCTGTTTTTTTTAGTTCTGTAACAATG
KU196374.1      CATATGTTTATGGTTGGTATGGATGTAAAGACTTCTGTTTTTTTTAGTTCTGTAACAATG
KU196341.1      CATATGTTTATGGTTGGTATGGATGTAAAGACTTCTGTTTTTTTTAGTTCTGTAACAATG
KU196321.1      CATATGTTTATGGTTGGTATGGATGTAAAGACTTCTGTTTTTTTTAGTTCTGTAACAATG
KU196301.1      CATATGTTTATGGTTGGTATGGATGTAAAGACTTCTGTTTTTTTTAGTTCTGTAACAATG
KU196331.1      CATATGTTTATGGTTGGTATGGATGTAAAGACTTCTGTTTTTTTTAGTTCTGTAACAATG
KU196322.1      CATATGTTTATGGTTGGTATGGATGTAAAGACTTCTGTTTTTTTTAGTTCTGTAACAATG
KU196312.1      CATATGTTTATGGTTGGTATGGATGTAAAGACTTCTGTTTTTTTTAGTTCTGTAACAATG
EU325878.1      CATATGTTTATGGTTGGTATGGATGTAAAGACTTCTGTTTTTTTTAGTTCTGTAACAATG
KU196395.1      CATATGTTTATGGTTGGTATGGATGTAAAGACTTCTGTTTTTTTTAGTTCTGTAACAATG
KF279409.1      CATATGTTTATGGTTGGTATGGATGTAAAGACTTCTGTTTTTTTTAGTTCTGTAACAATG
KF279408.1      CATATGTTTATGGTTGGTATGGATGTAAAGACTTCTGTTTTTTTTAGTTCTGTAACAATG
KF279407.1      CATATGTTTATGGTTGGTATGGATGTAAAGACTTCTGTTTTTTTTAGTTCTGTAACAATG
HM120847.1      CATATGTTTATGGTTGGTATGGATGTAAAGACTTCTGTTTTTTTTAGTTCTGTAACAATG
KR855674.1      CATATGTTTATGGTTGGTATGGATGTAAAGACTTCTGTTTTTTTTAGTTCTGTAACAATG
KU196417.1      CATATGTTTATGGTTGGTATGGATGTAAAGACTTCTGTTTTTTTTAGTTCTGTAACAATG
KU196413.1      CATATGTTTATGGTTGGTATGGATGTAAAGACTTCTGTTTTTTTTAGTTCTGTAACAATG
KU196367.1      CATATGTTTATGGTTGGTATGGATGTAAAGACTTCTGTTTTTTTTAGTTCTGTAACAATG
HM120848.1      CATATGTTTATGGTTGGTATGGATGTAAAGACTTCTGTTTTTTTTAGTTCTGTAACAATG
JQ781208.1      CATATGTTTATGGTTGGTATGGATGTAAAGACTTCTGTTTTTTTTAGTTCTGTAACAATG
JQ781209.1      CATATGTTTATGGTTGGTATGGATGTAAAGACTTCTGTTTTTTTTAGTTCTGTAACAATG
JQ781213.1      CATATGTTTATGGTTGGTATGGATGTAAAGACTTCTGTTTTTTTTAGTTCTGTAACAATG
JQ781212.1      CATATGTTTATGGTTGGTATGGATGTAAAGACTTCTGTTTTTTTTAGTTCTGTAACAATG
JQ781206.1      CATATGTTTATGGTTGGTATGGATGTAAAGACTTCTGTTTTTTTTAGTTCTGTAACAATG
KU196355.1      CATATGTTTATGGTTGGTATGGATGTAAAGACTTCTGTTTTTTTTAGTTCTGTAACAATG
JQ781210.1      CATATGTTTATGGTTGGTATGGATGTAAAGACTTCTGTTTTTTTTAGTTCTGTAACAATG
KU196388.1      CATATGTTTATGGTTGGTATGGATGTAAAGACTTCTGTTTTTTTTAGTTCTGTAACAATG
KU196379.1      CATATGTTTATGGTTGGTATGGATGTAAAGACTTCTGTTTTTTTTAGTTCTGTAACAATG
JQ781214.1      CATATGTTTATGGTTGGTATGGATGTAAAGACTTCTGTTTTTTTTAGTTCTGTAACAATG
KU196305.1      CATATGTTTATGGTTGGTATGGATGTAAAGACTTCTGTTTTTTTTAGTTCTGTAACAATG
                ************************************************************
```

**Supplementary Figure 1. MUSCLE alignment of *cox*1 nucleotide sequences from selected isolates of *Schistosoma japonicum*.** The nucleotide sequences accessed from GenBank and aligned here are detailed in Supplementary Table 2. The sequences were aligned using MUSCLE ^1^ with default parameters. Primer binding sites for the amplification of the 446-base target region are highlighted in grey. The 22-base target regions are also highlighted as follows: probe 2 target (yellow), probe 3 target (blue), probe 4 target (red) and probe 5 target (green). Only relevant portion of the MUSCLE alignment is shown.

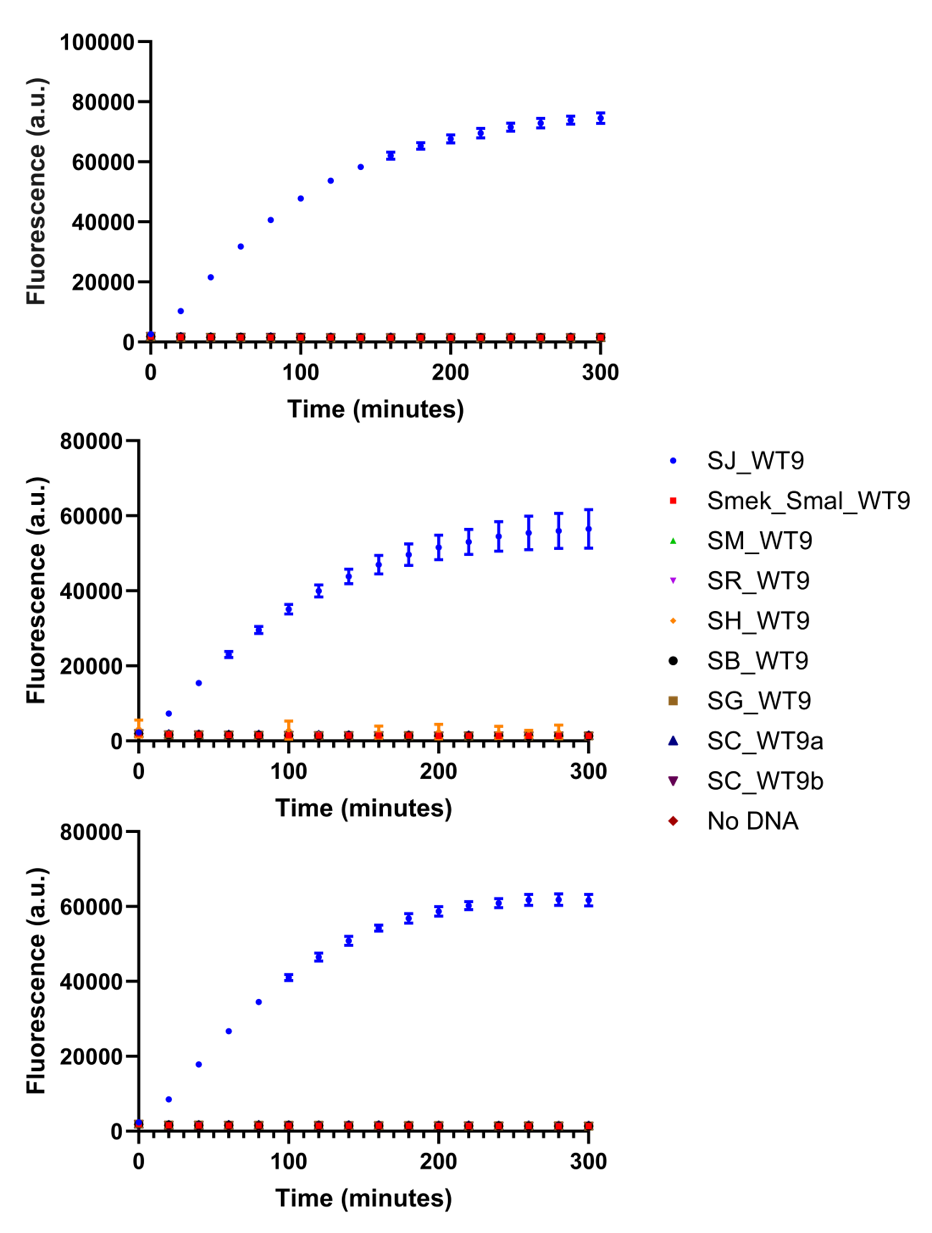

**Supplementary Figure 2. Specificity of *S. japonicum* probe set 2 against a range of DNA targets.** Both half probes (SJ_A2 and SJ_B2) and the target DNA concentrations were tested at 50 nM. Targets are listed in the key and further details of these targets and the probes are supplied in Supplementary Table 1. Three reaction runs are shown separately, with *n*=3 per graph (1 replicate per reaction, each reaction split into triplicate runs). Measurements were obtained using a BMG CLARIOstar plate reader (Ex. 440-15 nm/ Em. 510-20 nm, 1500 gain). Error bars denote standard error of the mean.

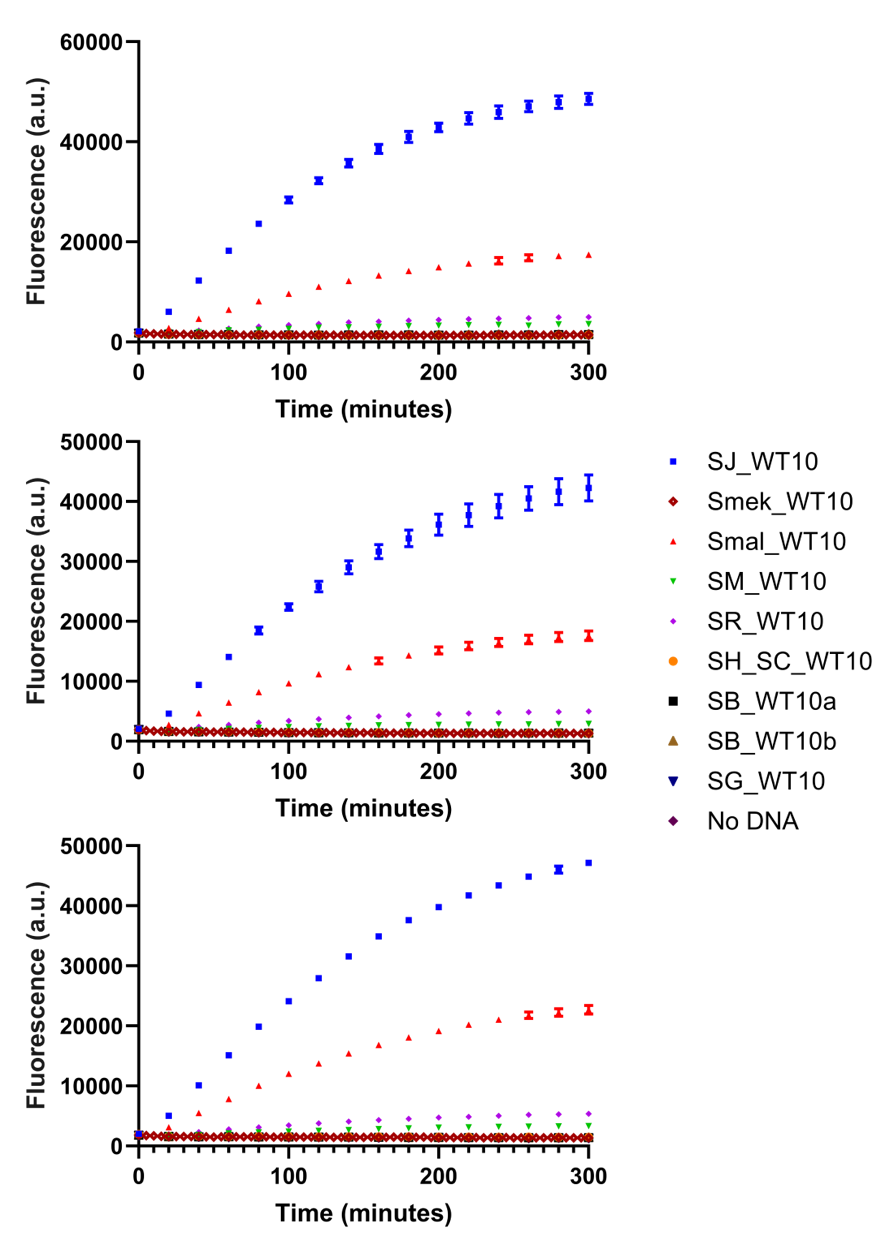

**Supplementary Figure 3. Specificity of *S. japonicum* probe set 3 against a range of DNA targets.** Both half probes (SJ_A3 and SJ_B3) and the target DNA concentrations were tested at 50 nM. Targets are listed in the key and further details of these targets and the probes are supplied in Supplementary Table 1. Three reaction runs are shown separately, with *n*=3 per graph (1 replicate per reaction, each reaction split into triplicate runs). Measurements were obtained using a BMG CLARIOstar plate reader (Ex. 440-15 nm/ Em. 510-20 nm, 1500 gain). Error bars denote standard error of the mean.

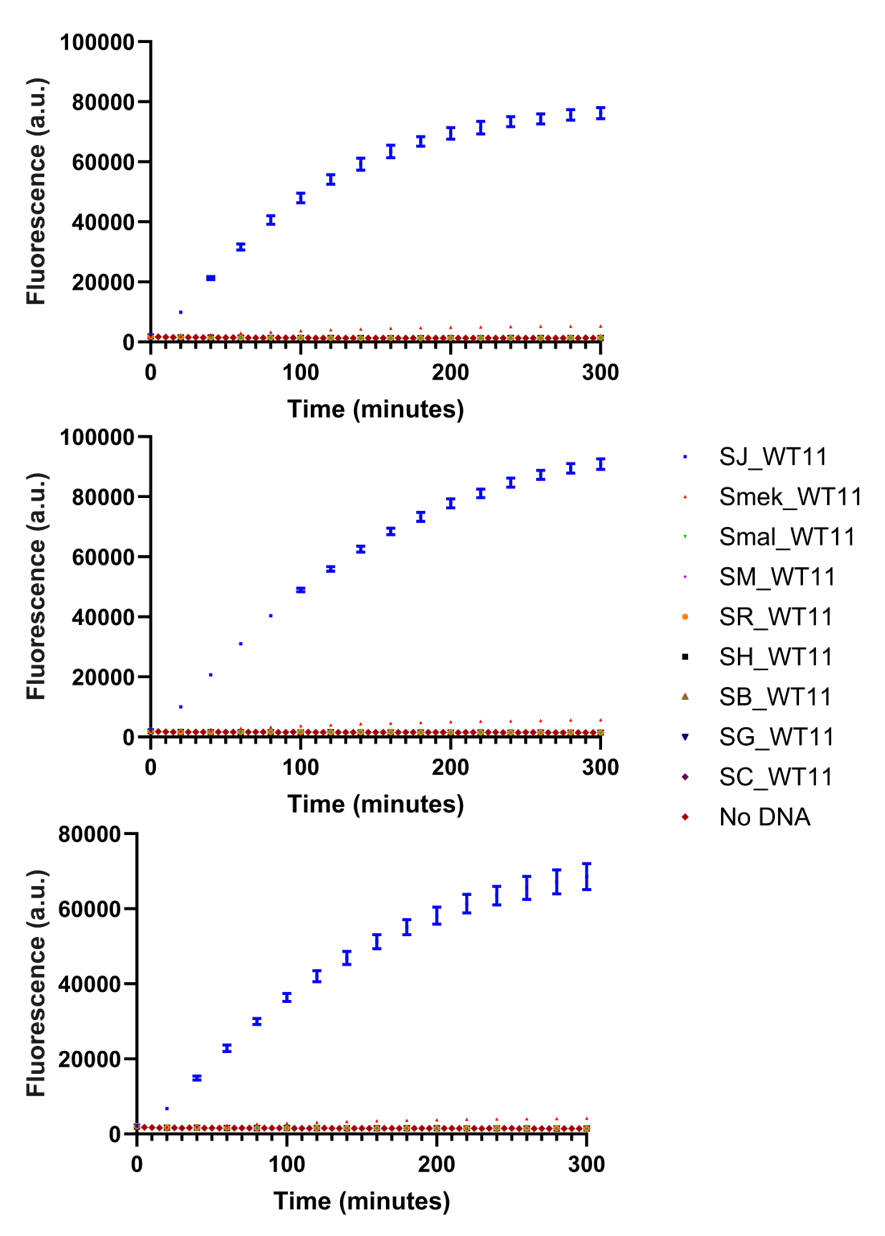

**Supplementary Figure 4. Specificity of *S. japonicum* probe set 4 against a range of DNA targets.** Both half probes (SJ_A4 and SJ_B4) and the target DNA concentrations were tested at 50 nM. Targets are listed in the key and further details of these targets and the probes are supplied in Supplementary Table 1. Three reaction runs are shown separately, with *n*=3 per graph (1 replicate per reaction, each reaction split into triplicate runs). Measurements were obtained using a BMG CLARIOstar plate reader (Ex. 440-15 nm/ Em. 510-20 nm, 1500 gain). Error bars denote standard error of the mean.

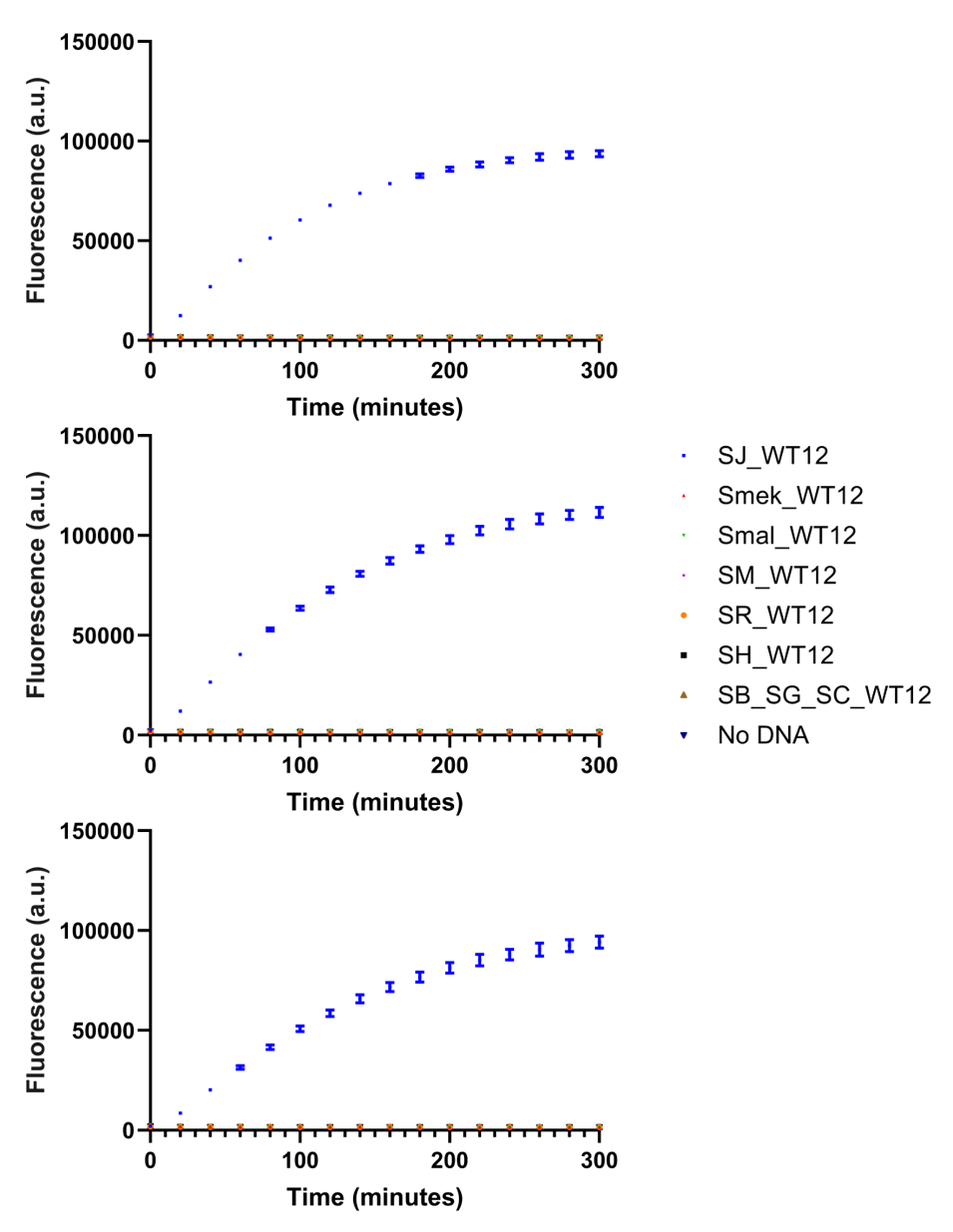

**Supplementary Figure 5. Specificity of *S. japonicum* probe set 5 against a range of DNA targets.** Both half probes (SJ_A5 and SJ_B5) and the target DNA concentrations were tested at 50 nM. Targets are listed in the key and further details of these targets and the probes are supplied in Supplementary Table 1. Three reaction runs are shown separately, with *n*=3 per graph (1 replicate per reaction, each reaction split into triplicate runs). Measurements were obtained using a BMG CLARIOstar plate reader (Ex. 440-15 nm/ Em. 510-20 nm, 1500 gain). Error bars denote standard error of the mean.

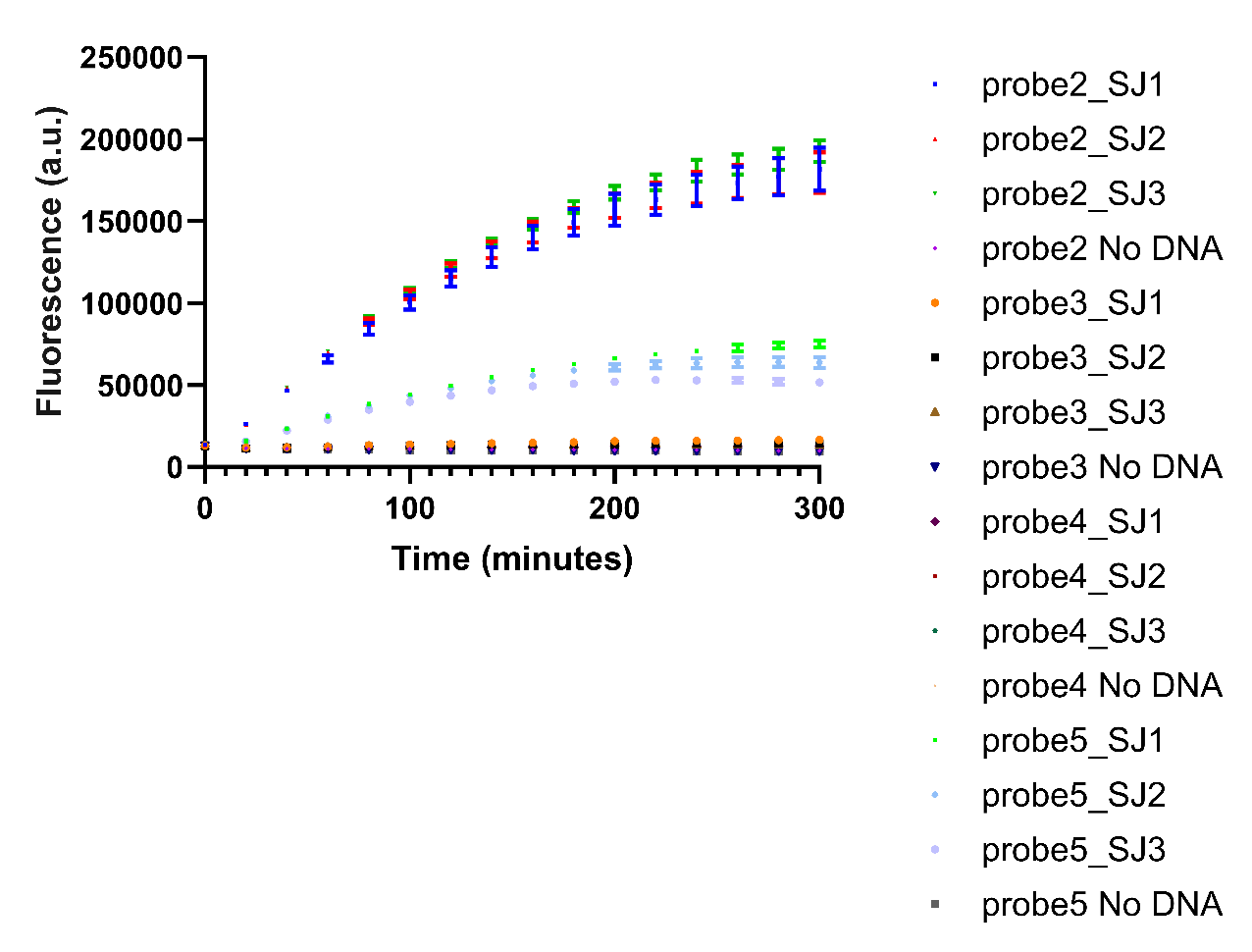

**Supplementary Figure 6. Recognition of ssDNA derived from plasmid DNA by *S. japonicum* probe sets 2, 3, 4 and 5.** The 446-base target region was PCR amplified from plasmid pAJW333 and treated as described in the main text to produce purified ssDNA. Half probes were tested at 50 nM and the target ssDNA tested at 30 ng. Three PCR reactions were tested (SJ1-SJ3) against each probe set. Reactions are identified as follows: probe2_SJ1-3, probe3_SJ1-3, probe4_SJ1-3 and probe5_SJ1-3 (*S. japonicum* probes 2, 3, 4 and 5 against PCR reactions 1-3 respectively), probe2 No DNA, probe3 No DNA, probe4 No DNA and probe5 No DNA (*S. japonicum* probes 2, 3, 4 and 5 negative controls respectively i.e. no target DNA). *n* =3 (1 replicate per each reaction split into triplicate runs). Measurements were obtained using a BMG CLARIOstar plate reader (Ex. 440-15 nm/ Em. 510-20 nm, 2000 gain). Error bars denote standard error of the mean.

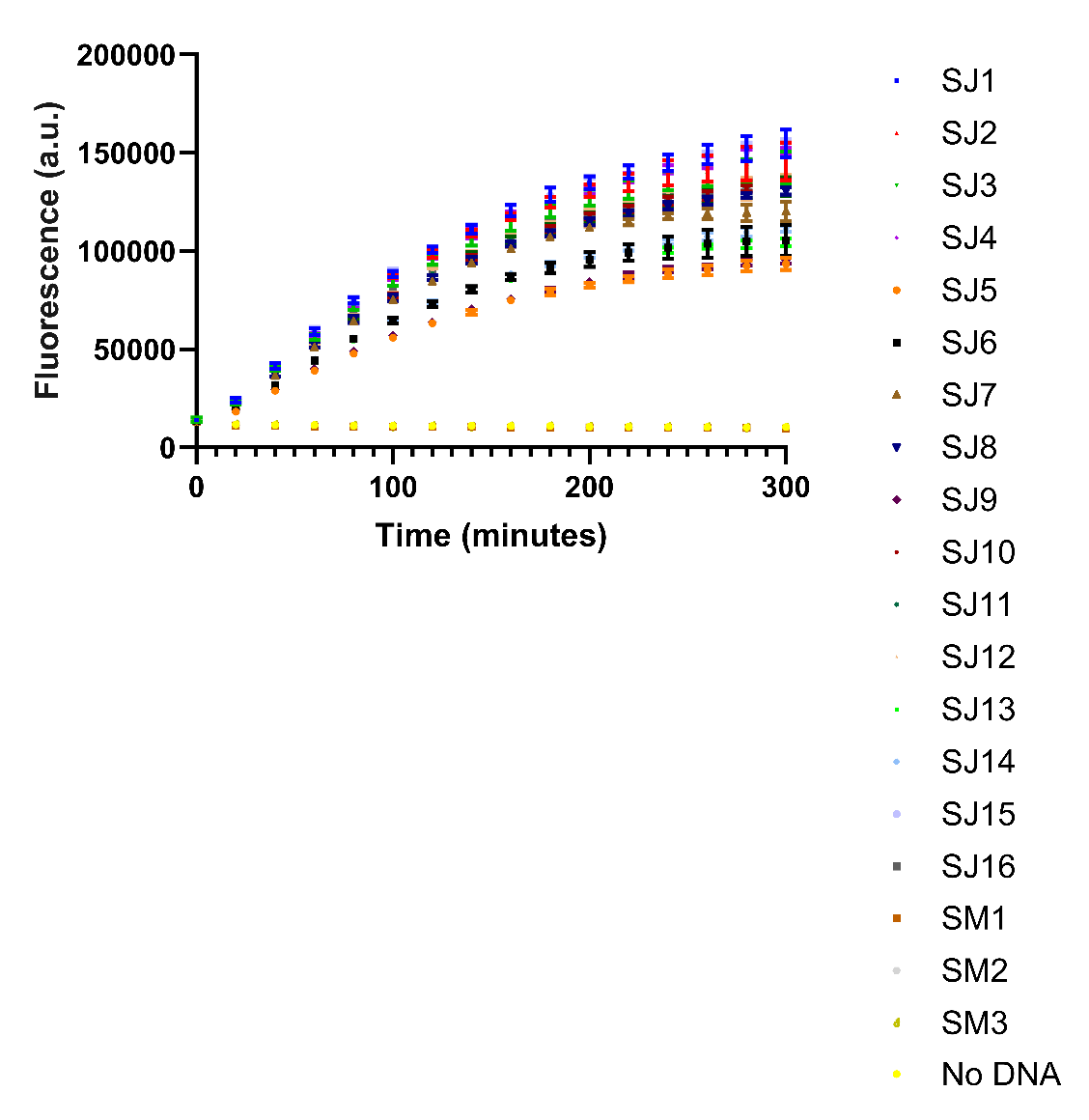

**Supplementary Figure 7. Probe 2 detection of ssDNA derived from *S. japonicum* and *S. mansoni* cercariae samples.** The 446-base target region was PCR amplified from the *Schistosoma* cercariae samples listed in Supplementary Table 6 and treated as described in the main text to produce purified ssDNA. Probe 2 half probes (SJ_A2 and SJ_B2) were tested at 50 nM and the target ssDNA tested at 30 ng. Reactions are identified as follows: SJ1-SJ16 relate to the ssDNA generated for *S. japonicum* cercariae samples 1-16 tested against probe 2, SM1-SM3 relate to the ssDNA generated for *S. mansoni* cercariae samples 1-3 against probe 2, No DNA is probe 2 negative control (no target ssDNA). *n* =3 (1 replicate per each reaction split into triplicate runs). Measurements were obtained using a BMG CLARIOstar plate reader (Ex. 440-15 nm/ Em. 510-20 nm, 2000 gain). Error bars denote standard error of the mean.

**Supplementary Table 1. Oligonucleotide probes and primers used in this study**

| **Number** | **Name** | **Sequence (5’-3’)** |
| --- | --- | --- |
| **Probes** | | |
| AJW694 | 2^nd^ T7 promoter sequence | **TAATACGACTCACTATAGGG** |
| AJW1011 | SJ_A2 | **p-ACATTATAGTAGCGATACCCTATAGTGAGTCGTATTA** |
| AJW1012 | SJ_B2 | **GTGTGGGAGCCCACACTCTACTCGACAGATACGAATATCTGGACCCGACCGTCTCCCACACCCTTAGACGCA** |
| AJW1013 | SJ_A3 | **p-ACTTCTGGGTGGCGATACCCTATAGTGAGTCGTATTA** |
| AJW1014 | SJ_B3 | **GTGTGGGAGCCCACACTCTACTCGACAGATACGAATATCTGGACCCGACCGTCTCCCACACTCAAAACATAT** |
| AJW1041 | SJ_A4 | **p-CTAATAACTGAGCGATACCCTATAGTGAGTCGTATTA** |
| AJW1042 | SJ_B4 | **GTGTGGGAGCCCACACTCTACTCGACAGATACGAATATCTGGACCCGACCGTCTCCCACACAATAAGATCAT** |
| AJW1043 | SJ_A5 | **p-CGAAGTAAATAGCGATACCCTATAGTGAGTCGTATTA** |
| AJW1044 | SJ_B5 | **GTGTGGGAGCCCACACTCTACTCGACAGATACGAATATCTGGACCCGACCGTCTCCCACACAATAACAACAC** |
| **Targets** | | |
| AJW1023 | SM_WT_9 | TACGATTTTTAGTCGTTTAAGA |
| AJW1024 | SR_WT_9 | TACTATTCTAAGTCGTTTAAGG |
| AJW1025 | SH_WT_9 | TACGATTATTAGTCGTGTCAAT |
| AJW1026 | SB_WT_9 | TACAATTATGAGTTGTGTAGAT |
| AJW1027 | SG_WT_9 | TACGATTATTAGTTGTGTTGAT |
| AJW1028 | SC_WT_9_AY157210 | TACAATTATTAGTCGTGTTGAT |
| AJW1029 | SC_WT_9_AJ519516 | TACAATTATCAGTCGTGTTGAT |
| AJW1030 | SJ_WT_9 | TACTATAATGTTGCGTCTAAGG |
| AJW1031 | Smek_Smal_WT_9 | TACAATAATGATACGTTTAAAT |
| AJW1032 | SM_WT_10 | CATCCAGAGGTTTATGTTTTGA |
| AJW1033 | SR_WT_10 | CATCCAGAAGTTTATGTTTTAA |
| AJW1034 | SH_SC_WT_10 | CATCCGGAGGTGTATGTTTTAA |
| AJW1035 | SB_WT_10_MH647124 | CATCCGGAGGTGTATGTTTTGA |
| AJW1036 | SB_WT_10_FJ897160 | CATCCTGAGGTGTATGTTTTGA |
| AJW1037 | SG_WT_10 | CATCCGGAGGTGTACGTTTTGA |
| AJW1038 | SJ_WT_10 | CACCCAGAAGTATATGTTTTGA |
| AJW1039 | Smek_WT_10 | CATCCTGAGGTTTATGTTTTAA |
| AJW1040 | Smal_WT_10 | CATCCTGAAGTTTATGTTTTGA |
| AJW1045 | SJ_WT_11 | TCAGTTATTAGATGATCTTATT |
| AJW1046 | Smek_WT_11 | TCAGTTATAAGTTGATCTTATT |
| AJW1047 | Smal_WT_11 | TCGGTTATAAGTTGGTCTTATT |
| AJW1048 | SM_WT_11 | TCGATAATAGTATGGGCTTATC |
| AJW1049 | SR_WT_11 | TCGATAATAGTGTGGGCTTATT |
| AJW1050 | SH_WT_11 | TCTATAATAATATGATCATATT |
| AJW1051 | SB_WT_11 | TCGATAATAATCTGATCATATT |
| AJW1052 | SG_WT_11 | TCAATAATAATTTGGTCATATT |
| AJW1053 | SC_WT_11 | TCAATAATAATCTGGTCATATT |
| AJW1054 | SJ_WT_12 | TATTTACTTCGGTGTTGTTATT |
| AJW1055 | Smek_WT_12 | TTTTTACTTCTATACTTTTGTT |
| AJW1056 | Smal_WT_12 | TTTTTACTTCTATACTTTTATT |
| AJW1057 | SM_WT_12 | TATTTACGTCTGTCTTACTATT |
| AJW1058 | SR_WT_12 | TATTTACATCTATTCTTTTATT |
| AJW1059 | SH_WT_12 | TGTTCACTTCTATCTTATTATT |
| AJW1060 | SB_SG_SC_WT_12 | TATTTACTTCAATTTTATTGTT |
| **Primers for target amplification and/or sequencing** | | |
| AJW1061 | 5-SJ-cox1 | CCTTTGTCTTCTTTAGCTACTTCTGG |
| AJW1062 | 3-SJ-cox1 | CTCCCCAAACACACAATAGAACCCA |
| AJW1063 | p-3-SJ-cox1 | p-CTCCCCAAACACACAATAGAACCCA |
| AJW1064 | 5-SM-cox1 | CCTTTATCAATTTGAGAGGGGTCTGG |
| AJW1065 | 3-SM-cox1 | CTACCTAAGCATACTATAGAAGCCA |
| AJW1065 | p-3-SM-cox1 | p-CTACCTAAGCATACTATAGAAGCCA |

The T7 promoter sequence, target sequences and the ‘Spinach’ aptamer sequence for the probes are indicated by red, blue and green text respectively. Phosphorylated primers are indicated by ‘p-’. The 2^nd^ T7 promoter primer sequence (AJW694) was taken from^2,3.^

**Supplementary Table 2. GenBank accession numbers of the *Schistosoma japonicum* *cox*1 gene sequences used to valid biosensor targets**

| **Strain/Isolate** | **Accession number** | **Location isolated** | **Reference** |
| --- | --- | --- | --- |
| Yunnan DaLi (SjYD) | EU325878.1 | China: Yunnan province | GenBank |
| SJLEYM5 | JQ781210.1 | Philippines: Leyte | GenBank |
| SJ7_pilon | KR855674.1 | China: Dali, Yunnan | ^4^ |
| YNEY10 | KU196417.1 | - | GenBank |
| YNEY06 | KU196413.1 | - | GenBank |
| JXDC09 | KU196367.1 | - | GenBank |
| SJYEIIF10 | HM120848.1 | China: Yunnan, Eryuan | GenBank |
| SCXC08 | KU196395.1 | - | GenBank |
| JXNC06 | KU196374.1 | - | GenBank |
| HNYY04 | KU196341.1 | - | GenBank |
| HBSS04 | KU196321.1 | - | GenBank |
| AHGC03 | KU196301.1 | - | GenBank |
| Yunnan Heqing | KF279409.1 | China: Yunnan, Heqing | GenBank |
| Yunnan Eryuan | KF279408.1 | China: Yunnan, Eryuan | GenBank |
| Sichuan Xichang | KF279407.1 | China: Sichuan, Xichang | GenBank |
| SJYEIIm10 | HM120847.1 | China: Yunnan, Eryuan | GenBank |
| HNYY06 | KU196343.1 | - | GenBank |
| HNCD04 | KU196331.1 | - | GenBank |
| AHTL10 | KU196317.1 | - | GenBank |
| AHTL05 | KU196312.1 | - | GenBank |
| AHGC07 | KU196305.1 | - | GenBank |
| PH0010 | KU196388.1 | - | GenBank |
| PH0001 | KU196379.1 | - | GenBank |
| IN0008 | KU196355.1 | - | GenBank |
| HBSS05 | KU196322.1 | - | GenBank |
| SJASNM6 | JQ781208.1 | Philippines: Asuncion | GenBank |
| SJLEYF5 | JQ781209.1 | Philippines: Leyte | GenBank |
| SJSXM24 | HM120846.1 | China: Sichuan, Tianquan | GenBank |
| SJSORM5 | JQ781214.1 | Philippines: Sorsogon | GenBank |
| SJSORF5 | JQ781213.1 | Philippines: Sorsogon | GenBank |
| SJMINM6 | JQ781212.1 | Philippines: Mindoro | GenBank |
| SJMINF5 | JQ781211.1 | Philippines: Mindoro | GenBank |
| SJASNF5 | JQ781206.1 | Philippines: Asuncion | GenBank |

**Supplementary Table 3. Bacterial strains and constructs used in this study.**

| **Strain or plasmid** | **Relevant features** | **Reference(s)** |
| --- | --- | --- |
| **Strain** |  |  |
| NEB10-beta | Δ(*ara -leu*) 7697 *araD139 fhuA* Δ*lacX74 galK16 galE15 e14-*ϕ*80*d*lacZ*Δ*M15 recA1 relA1endA1 nupG rpsL* (StrR) *rph spoT1* Δ(*mrr-hsdRMS-mcrBC*); cloning strain | New England Biolabs |
| **Plasmid** |  |  |
| pCR-Blunt-II-TOPO | Cloning vector for PCR products; KanR, ZeoR | Invitrogen |
| pAJW333 | NEB10-beta pCR-Blunt-II-TOPO-SJ-*cox*1; 446 base pair *cox*1 gene sequence fragment taken from GenBank entry EU325878.1, synthesised by IDT and cloned into pCR-II-TOPO; KanR, NeoR | This study |

**Supplementary Table 4. Δ fluorescence (a.u.) per hour measurements for *S. japonicum* probe sets 2, 3, 4 and 5 against corresponding target sequences from related species**

| **Probe and ssDNA target combination** | **Δ fluorescence (a.u.) per hour** | **Mean Δ fluorescence (a.u.) per hour** | **Standard error of the Mean** |
| --- | --- | --- | --- |
| Probe 2 SJ_WT9 | 30052, 29668, 31266, 22709, 21569, 22704, 25861, 25346, 26775 | 26217 | 1176 |
| Probe 2 Smek_Smal_WT9 | -95, -123, -101, -175, -159, -83, -84, -87, -85 | -110.2 | 11.58 |
| Probe 2 SM_WT9 | -137, -59, -152, -213, -116, -83, -93, -55, -63 | -107.9 | 17.46 |
| Probe 2 SR_WT9 | -97, -123, -143, -168, -143, -276, -71, -59, -95 | -130.6 | 21.72 |
| Probe 2 SH_WT9 | -99, -173, -124, -849, -158, -102, -111, -107, -97 | -202.2 | 81.34 |
| Probe 2 SB_WT9 | -70, -108, -175, -148, -210, -133, -114, -109, -75 | -126.9 | 15.13 |
| Probe 2 SG_WT9 | -143, -173, -184, -146, -94, -159, -58, -20, -40 | -113 | 20.45 |
| Probe 2 SC_WT9a | -95, -87, -126, -172, -200, -108, -53, -84, -9 | -103.8 | 19.27 |
| Probe 2 SC_WT9b | -103, -137, -141, -159, -159, -129, -37, -81, -100 | -116.2 | 13.35 |
| Probe 2 No DNA | -83, -111, -139, -119, -107, -180, -12, -75, -45 | -96.78 | 16.68 |
| Probe 3 SJ_WT10 | 17474, 17229, 18072, 13271, 13952, 14372, 14969, 14649, 14877 | 15429 | 571.2 |
| Probe 3 Smek_WT10 | -138, -160, -135, -186, -154, -151, -96, -39, -23 | -120.2 | 18.70 |
| Probe 3 Smal_WT10 | 5242, 5171, 5723, 5286, 5366, 5641, 6905, 6869, 6896 | 5900 | 254.6 |
| Probe 3 SM_WT10 | 123, 621, 615, 464, 444, 433, 581, 524, 545 | 483.3 | 50.77 |
| Probe 3 SR_WT10 | 1157, 1222, 1185, 1228, 1080, 1023, 1228, 1097, 1191 | 1157 | 24.65 |
| Probe 3 SH_SC_WT10 | -190, -134, -77, -140, -163, -119, -70, -4, -91 | -109.8 | 18.65 |
| Probe 3 SB_WT10a | -112, -93, -119, -203, -149, -111, -97, -63, -78 | -113.9 | 13.85 |
| Probe 3 SB_WT10b | -211, -145, -63, -150, -167, -113, -54, -27, -15 | -105 | 22.77 |
| Probe 3 SG_WT10 | -173, -132, -121, -133, -164, -172, -96, -47, -76 | -123.8 | 14.66 |
| Probe 3 No DNA | -133, -167, -110, -101, -146, -158, -83, -91, -76 | -118.3 | 11.23 |
| Probe 4 SJ_WT11 | 29789, 30117, 32290, 29589, 30443, 30860, 23575, 22512, 23468 | 28071 | 1253 |
| Probe 4 Smek_WT11 | 1320, 1365, 1403, 1267, 1333, 1325, 864, 774, 817 | 1163 | 87.36 |
| Probe 4 Smal_WT11 | -118, -175, -190, -109, -43, -149, -119, -91, -17 | -112.3 | 18.91 |
| Probe 4 SM_WT11 | -209, -116, -93, -63, -95, -90, -74, -57, -48 | -93.89 | 16.06 |
| Probe 4 SR_WT11 | -161, -248, -185, -84, -68, -52, -137, -85, -111 | -125.7 | 21.15 |
| Probe 4 SH_WT11 | -191, -192, -146, -51, -44, -60, -116, -84, -77 | -106.8 | 19.23 |
| Probe 4 SB_WT11 | -202, -162, -201, -73, -72, -85, -117, -111, -70 | -121.4 | 18.01 |
| Probe 4 SG_WT11 | -200, -178, -151, -92, -61, -44, -122, -59, -58 | -107.2 | 19.32 |
| Probe 4 SC_WT11 | -179, -190, -227, -79, -80, -49, -100, -60, -63 | -114.1 | 22.08 |
| Probe 4 No DNA | -173, -150, -169, -19, -42, -76, -75, -22, -110 | -92.89 | 20.22 |
| Probe 5 SJ_WT12 | 38931, 39002, 38982, 40116, 41309, 41011, 32083, 33970, 32993 | 37600 | 1191 |
| Probe 5 Smek_WT12 | -185, -152, -189, -104, -44, -141, -87, -71, -84 | -117.4 | 17.16 |
| Probe 5 Smal_WT12 | -177, -127, -192, -125, -67, -113, -86, -106, -121 | -123.8 | 13.22 |
| Probe 5 SM_WT12 | -166, -194, -168, -40, -130, -55, -98, -89, -124 | -118.2 | 17.50 |
| Probe 5 SR_WT12 | -160, -173, -185, -142, -89, -4, -15, 2, -113 | -97.67 | 25 |
| Probe 5 SH_WT12 | -119, -175, -165, 5, -30, -25, -58, -100, -61 | -80.89 | 21 |
| Probe 5 SB_SG_SC_WT12 | -165, -178, -130, -77, -106, -36, -92, -21, -39 | -93.78 | 18.84 |
| Probe 5 No DNA | -157, -187, -177, -44, -31, -29, -74, -49, -86 | -92.67 | 21.30 |

Data is shown in Fig. 1 of the main manuscript. Δ fluorescence (a.u.) per hour was calculated using raw fluorescence values between 20 and 80 minutes of the plate reader assay. Measurements were obtained using a BMG CLARIOstar plate reader (Ex. 440-15 nm/ Em. 510-20 nm, 1500 gain). Mean and Standard error of the mean calculated using GraphPad Prism 10.4.1.

**Supplementary Table 5. Δ fluorescence (a.u.) per hour measurements for *S. japonicum* probe sets 2, 3, 4 and 5 against ssDNA derived from plasmids containing the 446-base *S. japonicum*-specific *cox*1 target region**

| **Probe and ssDNA target combination** | **Δ fluorescence (a.u.) per hour** | **Mean Δ fluorescence (a.u.) per hour** | **Standard error of the Mean** |
| --- | --- | --- | --- |
| probe2_SJ1 | 55433, 56645, 62301 | 58126 | 2116 |
| probe2_SJ2 | 61212, 62606, 65333 | 63050 | 1210 |
| probe2_SJ3 | 62594, 65502, 65250 | 64449 | 930.2 |
| probe2 No DNA | -421, -464, -479 | -454.7 | 17.38 |
| probe3_SJ1 | 1451, 1668, 1373 | 1497 | 88.25 |
| probe3_SJ2 | 946, 1263, 1381 | 1197 | 129.9 |
| probe3_SJ3 | 1634, 1319, 848 | 1267 | 228.4 |
| probe3 No DNA | -744, -408, -535 | -562.3 | 97.95 |
| probe4_SJ1 | 744, 850, 297 | 630.3 | 169.5 |
| probe4_SJ2 | 254, 10, 1105 | 456.3 | 331.9 |
| probe4_SJ3 | 83, 241, 938 | 420.7 | 262.7 |
| probe4 No DNA | -616, -670, -360 | -548.7 | 95.61 |
| probe5_SJ1 | 22705, 22901, 22359 | 22655 | 158.4 |
| probe5_SJ2 | 21986, 21167, 21794 | 21649 | 247.3 |
| probe5_SJ3 | 19456, 19341, 19296 | 19364 | 47.64 |
| probe5 No DNA | -758, -547, -618 | -641.0 | 61.99 |

Data is shown in Fig. 2 of the main manuscript. Δ fluorescence (a.u.) per hour was calculated using raw fluorescence values between 20 and 80 minutes of the plate reader assay. Measurements were obtained using a BMG CLARIOstar plate reader (Ex. 440-15 nm/ Em. 510-20 nm, 2000 gain). Mean and Standard error of the mean calculated using GraphPad Prism 10.4.1.

**Supplementary Table 6. Sequences of *cox*1 amplified from either *S. japonicum* or *S. mansoni*** **cercarial gDNA**

| **Sample number** | **Notes** | **Sequence** |
| --- | --- | --- |
| SJ1 | *S. japonicum*, Tongqiao  OZ203289 | TTTAGCTACTTCTGGTGTTGGTGTGGATTACTTAATGTTCTCTTTACATCTTGCTGGTGTATCTAGTTTGATTGGTTCTATAAATTTTATTACTACTATAATGTTGCGTCTAAGGTCATGTTCTTCAGTTATTAGATGATCTTATTTATTTACTTCGGTGTTGTTATTGTTATCGTTGCCGGTTCTTGCTGCAGGTATAACTATGTTGTTGTTTGATCGTAAATTTGGTACTGCTTTTTTTGAGCCAGCAGGTGGTGGTGATCCTGTGTTATTTCAACATTTATTTTGGTTTTTTGGTCACCCAGAAGTATATGTTTTGATATTGCCTGGATTTGGTATAGTAAGTCATATATGTATGTCTTTAAGTAATAATAATTCTTCGTTTGGATATTATGGGTTAGTTTGTGCCATGGGTTCTATTGTG |
| SJ2 | *S. japonicum*, Tongqiao  OZ203290 | TTTAGCTACTTCTGGTGTTGGTGTGGATTACTTAATGTTCTCTTTACATCTTGCTGGTGTATCTAGTTTGATTGGTTCTATAAATTTTATTACTACTATAATGTTGCGTCTAAGGTCATGTTCTTCAGTTATTAGATGATCTTATTTATTTACTTCGGTGTTGTTATTGTTATCGTTGCCGGTTCTTGCTGCAGGTATAACTATGTTGTTGTTTGATCGTAAATTTGGTACTGCTTTTTTTGAGCCAGCAGGTGGTGGTGATCCTGTGTTATTTCAACATTTATTTTGGTTTTTTGGTCACCCAGAAGTATATGTTTTGATATTGCCTGGATTTGGTATAGTAAGTCATATATGTATGTCTTTAAGTAATAATAATTCTTCGTTTGGATATTATGGGTTAGTTTGTGCCATGGGTTCTATTGTGTGTTTGGGGAG |
| SJ3 | *S. japonicum*, Chenbi  OZ203291 | TTAGCTACTTCTGGTGTTGGTGTGGATTACTTAATGTTCTCTTTACATCTTGCTGGTGTATCTAGTTTGATTGGTTCTATAAATTTTATTACTACTATAATGTTGCGTCTAAGGTCATGTTCTTCAGTTATTAGATGATCTTATTTATTTACTTCGGTGTTGTTATTGTTATCGTTGCCGGTTCTTGCTGCAGGTATAACTATGTTGTTGTTTGATCGTAAATTTGGTACTGCTTTTTTTGAGCCAGCAGGTGGTGGTGATCCTGTGTTATTTCAACATTTATTTTGGTTTTTTGGTCACCCAGAAGTATATGTTTTGATATTGCCTGGATTTGGTATAGTAAGTCATATATGTATGTCTTTAAGTAATAATAATTCTTCGTTTGGATATTATGGGTTAGTTTGTGCCATGGGTTCTATTGTGTGTT |
| SJ4 | *S. japonicum*, Chenbi  OZ203292 | TTCTTTAGCTACTTCTGGTGTTGGTGTGGATTACTTAATGTTCTCTTTACATCTTGCTGGTGTATCTAGTTTGATTGGTTCTATAAATTTTATTACTACTATAATGTTGCGTCTAAGGTCATGTTCTTCAGTTATTAGATGATCTTATTTATTTACTTCGGTGTTGTTATTGTTATCGTTGCCGGTTCTTGCTGCAGGTATAACTATGTTGTTGTTTGATCGTAAATTTGGTACTGCTTTTTTTGAGCCAGCAGGTGGTGGTGATCCTGTGTTATTTCAACATTTATTTTGGTTTTTTGGTCACCCAGAAGTATATGTTTTGATATTGCCTGGATTTGGTATAGTAAGTCATATATGTATGTCTTTAAGTAATAATAATTCTTCGTTTGGATATTATGGGTTAGTTTGTGCCATGGGTTCTATTGTGTGTTTGGGGAG |
| SJ5 | *S. japonicum*, Chenbi  OZ203293 | CTTTGTCTTCTTTAGCTACTTCTGGTGTTGGTGTAGATTACTTAATGTTCTCTTTACATCTTGCTGGTGTATCTAGTTTGATTGGTTCTATAAATTTTATTACTACTATAATGTTGCGTCTAAGGTCATGTTCTTCAGTTATTAGATGATCTTATTTATTTACTTCGGTGTTGTTACTGTTATCGTTGCCGGTTCTTGCTGCAGGTATAACTATGTTGTTGTTTGATCGTAAATTTGGTACTGCTTTTTTTGAGCCAGCAGGTGGTGGTGATCCTGTGTTATTTCAACATTTATTTTGGTTTTTTGGTCACCCAGAAGTATATGTTTTGATATTGCCTGGATTTGGTATAGTAAGTCATATATGTATGTCTTTAAGTAATAATAATTCTTCGTTTGGATATTATGGGTTAGTTTGTGCTATGGGTTCTATTGTGTGT |
| SJ6 | *S. japonicum*, Chenbi  OZ203294 | CTTCTTTAGCTACTTCTGGTGTTGGTGTAGATTACTTAATGTTCTCTTTACATCTTGCTGGTGTATCTAGTTTGATTGGTTCTATAAATTTTATTACTACTATAATGTTGCGTCTAAGGTCATGTTCTTCAGTTATTAGATGATCTTATTTATTTACTTCGGTGTTGTTACTGTTATCGTTGCCGGTTCTTGCTGCAGGTATAACTATGTTGTTGTTTGATCGTAAATTTGGTACTGCTTTTTTTGAGCCAGCAGGTGGTGGTGATCCTGTGTTATTTCAACATTTATTTTGGTTTTTTGGTCACCCAGAAGTATATGTTTTGATATTGCCTGGATTTGGTATAGTAAGTCATATATGTATGTCTTTAAGTAATAATAATTCTTCGTTTGGATATTATGGGTTAGTTTGTGCTATGGGTTCTATTGTGTGTTT |
| SJ7 | *S. japonicum*, Jianhong  OZ203295 | TTTAGCTACTTCTGGTGTTGGTGTGGATTATTTAATGTTCTCTTTACATCTTGCTGGTGTATCTAGTTTGATTGGTTCTATAAATTTTATTACTACTATAATGTTGCGTCTAAGGTCATGTTCTTCAGTTATTAGATGATCTTATTTATTTACTTCGGTGTTGTTACTGTTATCGTTGCCGGTTCTTGCTGCAGGTATAACTATGTTGTTGTTTGATCGTAAATTTGGTACTGCTTTTTTTGAGCCAGCAGGTGGTGGTGATCCTGTGTTATTTCAACATTTATTTTGGTTTTTTGGTCACCCAGAAGTATATGTTTTGATATTGCCTGGATTTGGTATAGTAAGTCATATATGTATGTCTTTAAGTAATAATAATTCTTCGTTTGGATATTATGGGTTAGTTTGTGCTATGGGTTCTATTGTG |
| SJ8 | *S. japonicum*, Jianhong  OZ203296 | TTCTTTAGCTACTTCTGGTGTTGGTGTGGATTATTTAATGTTCTCTTTACATCTTGCTGGTGTATCTAGTTTGATTGGTTCTATAAATTTTATTACTACTATAATGTTGCGTCTAAGGTCATGTTCTTCAGTTATTAGATGATCTTATTTATTTACTTCGGTGTTGTTACTGTTATCGTTGCCGGTTCTTGCTGCAGGTATAACTATGTTGTTGTTTGATCGTAAATTTGGTACTGCTTTTTTTGAGCCAGCAGGTGGTGGTGATCCTGTGTTATTTCAACATTTATTTTGGTTTTTTGGTCACCCAGAAGTATATGTTTTGATATTGCCTGGATTTGGTATAGTAAGTCATATATGTATGTCTTTAAGTAATAATAATTCTTCGTTTGGATATTATGGGTTAGTTTGTGCTATGGGTTCTATTGTGTGTTT |
| SJ9 | *S. japonicum*, Jianhong  OZ203297 | TTCTTTAGCTACTTCTGGTGTTGGTGTGGATTATTTAATGTTCTCTTTACATCTTGCTGGTGTATCTAGTTTGATTGGTTCTATAAATTTTATTACTACTATAATGTTGCGTCTAAGGTCATGTTCTTCAGTTATTAGATGATCTTATTTATTTACTTCGGTGTTGTTACTGTTATCGTTGCCGGTTCTTGCTGCAGGTATAACTATGTTGTTGTTTGATCGTAAATTTGGTACTGCTTTTTTTGAGCCAGCAGGTGGTGGTGATCCTGTGTTATTTCAACATTTATTTTGGTTTTTTGGTCACCCAGAAGTATATGTTTTGATATTGCCTGGATTTGGTATAGTAAGTCATATATGTATGTCTTTAAGTAATAATAATTCTTCGTTTGGATATTATGGGTTAGTTTGTGCTATGGGTTCTATTGTGTGTTT |
| SJ10 | *S. japonicum*, Shundi  OZ203298 | CCTTTGTCTTCTTTAGCTACTTCTGGTGTTGGTGTGGATTACTTAATGTTCTCTTTACATCTTGCTGGTGTATCTAGTTTGATTGGTTCTATAAATTTTATTACTACTATAATGTTGCGTCTAAGGTCATGTTCTTCAGTTATTAGATGATCTTATTTATTTACTTCGGTGTTGTTATTGTTATCATTGCCGGTTCTTGCTGCAGGTATAACTATGTTGCTGTTTGATCGTAAATTTGGTACTGCTTTTTTTGAGCCAGCAGGTGGTGGTGATCCTGTGTTATTTCAACATTTATTTTGGTTTTTTGGTCACCCAGAAGTATATGTTTTGATATTGCCTGGATTTGGTATAGTAAGTCATATATGTATGTCTTTAAGTAATAATAATTCTTCGTTCGGATATTATGGGTTAGTTTGTGCTATGGGTTCTATTGTGTGTTT |
| SJ11 | *S. japonicum*, Shundi  OZ203299 | CCTTTGTCTTCTTTAGCTACTTCTGGTGTTGGTGTGGATTACTTAATGTTCTCTTTACATCTTGCTGGTGTATCTAGTTTGATTGGTTCTATAAATTTTATTACTACTATAATGTTGCGTCTAAGGTCATGTTCTTCAGTTATTAGATGATCTTATTTATTTACTTCGGTGTTGTTATTGTTATCGTTGCCGGTTCTTGCTGCAGGTATAACTATGTTGTTGTTTGATCGTAAATTTGGTACTGCTTTTTTTGAGCCAGCAGGTGGTGGTGATCCTGTGTTATTTCAACATTTATTTTGGTTTTTTGGTCACCCAGAAGTATATGTTTTGATATTGCCTGGATTTGGTATAGTAAGTCATATATGTATGTCTTTAAGTAATAATAATTCTTCGTTTGGATATTATGGGTTAGTTTGTGCCATGGGTTCTATTGTGTGTTTGGGGAG |
| SJ12 | *S. japonicum*, Shundi  OZ203300 | TTCTTTAGCTACTTCTGGTGTTGGTGTGGATTACTTAATGTTCTCTTTACATCTTGCTGGTGTATCTAGTTTGATTGGTTCTATAAATTTTATTACTACTATAATGTTGCGTCTAAGGTCATGTTCTTCAGTTATTAGATGATCTTATTTATTTACTTCGGTGTTGTTATTGTTATCGTTGCCGGTTCTTGCTGCAGGTATAACTATGTTGTTGTTTGATCGTAAATTTGGTACTGCTTTTTTTGAGCCAGCAGGTGGTGGTGATCCTGTGTTATTTCAACATTTATTTTGGTTTTTTGGTCACCCAGAAGTATATGTTTTGATATTGCCTGGATTTGGTATAGTAAGTCATATATGTATGTCTTTAAGTAATAATAATTCTTCGTTTGGATATTATGGGTTAGTTTGTGCCATGGGTTCTATTGTGTGTTT |
| SJ13 | *S. japonicum*, Shundi  OZ203301 | TTCTTTAGCTACTTCTGGTGTTGGTGTGGATTACTTAATGTTCTCTTTACATCTTGCTGGTGTATCTAGTTTGATTGGTTCTATAAATTTTATTACTACTATAATGTTGCGTCTAAGGTCATGTTCTTCAGTTATTAGATGATCTTATTTATTTACTTCGGTGTTATTATTGTTATCGTTGCCGGTTCTTGCTGCAGGTATAACTATGTTGTTGTTTGATCGTAAATTTGGTACTGCTTTTTTTGAGCCAGCAGGTGGTGGTGATCCTGTGTTATTTCAACATTTATTTTGGTTTTTTGGTCACCCAGAAGTATATGTTTTGATATTGCCTGGATTTGGTATAGTAAGTCATATATGTATGTCTTTAAGTAATAATAATTCTTCGTTTGGATATTATGGGTTAGTTTGTGCTATGGGTTCTATTGTGTGTTT |
| SJ14 | *S. japonicum*, Shundi  OZ203302 | TTCTTTAGCTACTTCTGGTGTTGGTGTGGATTACTTAATGTTCTCTTTACATCTTGCTGGTGTATCTAGTTTGATTGGTTCTATAAATTTTATTACTACTATAATGTTGCGTCTAAGGTCATGTTCTTCAGTTATTAGATGATCTTATTTATTTACTTCGGTGTTATTATTGTTATCGTTGCCGGTTCTTGCTGCAGGTATAACTATGTTGTTGTTTGATCGTAAATTTGGTACTGCTTTTTTTGAGCCAGCAGGTGGTGGTGATCCTGTGTTATTTCAACATTTATTTTGGTTTTTTGGTCACCCAGAAGTATATGTTTTGATATTGCCTGGATTTGGTATAGTAAGTCATATATGTATGTCTTTAAGTAATAATAATTCTTCGTTTGGATATTATGGGTTAGTTTGTGCTATGGGTTCTATTGTGTGTTTGGGGAG |
| SJ15 | *S. japonicum*, Shundi  OZ203303 | CTTCTTTAGCTACTTCTGGTGTTGGTGTAGATTACTTAATGTTCTCTTTACATCTTGCTGGTGTATCTAGTTTGATTGGTTCTATAAATTTTATTACTACTATAATGTTGCGTCTAAGGTCATGTTCTTCAGTTATTAGATGATCTTATTTATTTACTTCGGTGTTGTTATTGTTATCGTTGCCGGTTCTTGCTGCAGGTATAACTATGTTGTTGTTTGATCGTAAATTTGGTACTGCTTTTTTTGAGCCAGCAGGTGGTGGTGATCCTGTGTTATTTCAACATTTATTTTGGTTTTTTGGTCACCCAGAAGTATATGTTTTGATATTGCCTGGATTTGGTATAGTAAGTCATATATGTATGTCTTTAAGTAATAATAATTCTTCGTTTGGATATTATGGGTTGGTTTGTGCTATGGGTTCTATTGTGTGTTT |
| SJ16 | *S. japonicum*, Shundi  OZ203304 | CTTCTTTAGCTACTTCTGGTGTTGGTGTAGATTACTTAATGTTCTCTTTACATCTTGCTGGTGTATCTAGTTTGATTGGTTCTATAAATTTTATTACTACTATAATGTTGCGTCTAAGGTCATGTTCTTCAGTTATTAGATGATCTTATTTATTTACTTCGGTGTTGTTATTGTTATCGTTGCCGGTTCTTGCTGCAGGTATAACTATGTTGTTGTTTGATCGTAAATTTGGTACTGCTTTTTTTGAGCCAGCAGGTGGTGGTGATCCTGTGTTATTTCAACATTTATTTTGGTTTTTTGGTCACCCAGAAGTATATGTTTTGATATTGCCTGGATTTGGTATAGTAAGTCATATATGTATGTCTTTAAGTAATAATAATTCTTCGTTTGGATATTATGGGTTGGTTTGTGCTATGGGTTCTATTGTGTGTTT |
| SM1 | *S. mansoni*, laboratory-maintained strain | ATTTGAGAGGGGTCTGGTTTTGGTGTAGATTATTTAATGTTTTCTCTTCATTTGGCAGGGGTTTCAAGTCTAATTGGATCTGTCAATTTCATTTCTACGATTTTTAGTCGTTTAAGATTCAAATGTTCGATAATAGTATGGGCTTATCTATTTACGTCTGTTTTATTATTGCTTTCGTTACCTGTGTTAGCCAGAGGAATAACGATGTTATTATTTGATCGTAAATTTGGAACTGCTTTTTTTGAGCCGTCAGGCGGTGGCGATCCTATTTTGTTTCAGCATTTATTTTGGTTTTTTGGTCATCCAGAGGTTTATGTTTTGATCCTTCCGGGTTTTGGTATAGTTAGGCATATCTGTATGAGTCTAAGGAATAAAGATTCGTCGTTTGGTTATTATGGATTGATTTGCGCTATGGCTTCTATAGTATGC |
| SM2 | *S. mansoni*, laboratory-maintained strain | CAATTTGAGAGGGGTCTGGTTTTGGTGTAGATTATTTAATGTTTTCTCTTCATTTGGCAGGGGTTTCAAGTCTAATTGGATCTGTCAATTTCATTTCTACGATTTTTAGTCGTTTAAGATTCAAATGTTCGATAATAGTATGGGCTTATCTATTTACGTCTGTTTTATTATTGCTTTCGTTACCTGTGTTAGCCAGAGGAATAACGATGTTATTATTTGATCGTAAATTTGGAACTGCTTTTTTTGAGCCGTCAGGCGGTGGCGATCCTATTTTGTTTCAGCATTTATTTTGGTTTTTTGGTCATCCAGAGGTTTATGTTTTGATCCTTCCGGGTTTTGGTATAGTTAGGCATATCTGTATGAGTCTAAGGAATAAAGATTCGTCGTTTGGTTATTATGGATTGATTTGCGCTATGGCTTCTATAGTATGC |
| SM3 | *S. mansoni*, laboratory-maintained strain | CAATTTGAGAGGGGTCTGGTTTTGGTGTAGATTATTTAATGTTTTCTCTTCATTTGGCAGGGGTTTCAAGTCTAATTGGATCTGTCAATTTCATTTCTACGATTTTTAGTCGTTTAAGATTCAAATGTTCGATAATAGTATGGGCTTATCTATTTACGTCTGTTTTATTATTGCTTTCGTTACCTGTGTTAGCCAGAGGAATAACGATGTTATTATTTGATCGTAAATTTGGAACTGCTTTTTTTGAGCCGTCAGGCGGTGGCGATCCTATTTTGTTTCAGCATTTATTTTGGTTTTTTGGTCATCCAGAGGTTTATGTTTTGATCCTTCCGGGTTTTGGTATAGTTAGGCATATCTGTATGAGTCTAAGGAATAAAGATTCGTCGTTTGGTTATTATGGATTGATTTGCGCTATGGCTTCTATAGTATGC |

Bases highlighted in red indicate sequence variation from that of *S. japonicum* GenBank entry EU325878.1.

**Supplementary Table 7. Δ fluorescence (a.u.) per hour measurements for *S. japonicum*-specific probe 2 against ssDNA derived from cercariae samples SJ1-SJ16 and SM1-SM3**

| **ssDNA target** | **Δ fluorescence (a.u.) per hour** | **Mean Δ fluorescence (a.u.) per hour** | **Standard error of the Mean** |
| --- | --- | --- | --- |
| SJ1 | 51420, 50232, 50596 | 50749 | 351.4 |
| SJ2 | 50058, 50885, 50015 | 50319 | 283.1 |
| SJ3 | 48452, 48307, 49962 | 48907 | 529.2 |
| SJ4 | 49204, 49896, 50492 | 49864 | 372.2 |
| SJ5 | 28840, 29739, 30003 | 29527 | 352 |
| SJ6 | 36377, 35920, 34892 | 35730 | 439.1 |
| SJ7 | 42633, 42475, 43810 | 42973 | 421.1 |
| SJ8 | 41886, 42872, 44156 | 42971 | 657.2 |
| SJ9 | 29800, 30121, 30191 | 30037 | 120.4 |
| SJ10 | 44306, 43837, 43464 | 43869 | 243.6 |
| SJ11 | 44718, 44307, 45907 | 44977 | 479.7 |
| SJ12 | 42736, 45011, 45246 | 44331 | 800.4 |
| SJ13 | 35053, 34802, 35441 | 35099 | 185.9 |
| SJ14 | 36850, 35929, 36735 | 36505 | 289.7 |
| SJ15 | 52032, 53062, 49416 | 51503 | 1085 |
| SJ16 | 44342, 45292, 44363 | 44666 | 313.2 |
| SM1 | -635, -465, -517 | -539 | 50.29 |
| SM2 | -686, -231, -494 | -470.3 | 131.9 |
| SM3 | -239, -718, -294 | -417 | 151.3 |
| No DNA | -702, -662, -673 | -679 | 11.93 |

Data is shown in Fig. 3 of the main manuscript. Δ fluorescence (a.u.) per hour was calculated using raw fluorescence values between 20 and 80 minutes of the plate reader assay. Measurements were obtained using a BMG CLARIOstar plate reader (Ex. 440-15 nm/ Em. 510-20 nm, 2000 gain). Mean and Standard error of the mean calculated using GraphPad Prism 10.4.1.
